# Predicting individual long-term prognosis of spatial neglect based on acute stroke patient data

**DOI:** 10.1101/2024.01.10.24301050

**Authors:** Lisa Röhrig, Daniel Wiesen, Dongyun Li, Hans-Otto Karnath

## Abstract

One of the most pressing questions after a stroke is whether an individual patient will recover in the long-term. Previous studies demonstrated that spatial neglect – a common behavioral deficit after right hemispheric stroke – is a strong predictor for poor performance on a wide range of everyday tasks and for resistance to rehabilitation. The possibility of predicting long-term prognosis of spatial neglect is therefore of great relevance. The aim of the present study was to test the prognostic value of different imaging and non-imaging features from right hemispheric stroke patients: individual demographics (age, sex), initial neglect severity, and acute lesion information (size, location). Patients’ behavior was tested twice in the acute and the chronic phases of stroke and prediction models were built using machine learning-based algorithms with repeated nested cross-validation and feature selection. Model performances indicate that demographic information seemed less beneficial. The best variable combination comprised individual neglect severity in the acute phase of stroke, together with lesion location and size. The latter were based on individual lesion overlaps with a previously proposed chronic neglect region-of-interest (ROI) that covers anterior parts of the superior and middle temporal gyri and the basal ganglia. These variables achieved a remarkably high level of accuracy by explaining 66% of the total variance of neglect patients, making them promising features in the prediction of individual outcome prognosis.

## Introduction

Spatial neglect is one of the most frequent cognitive disorders following right hemisphere brain damage, forming the counterpart to aphasia after left hemisphere lesions. It is characterized by impaired orienting towards the contralesional (usually left) side, leading to neglect of contralesionally located objects or people. In contrast to stroke survivors without neglect, patients with spatial neglect experience prolonged inpatient periods, impaired functional recovery and a poor rehabilitation outcome if left untreated (Di Monaco et al., 2011; Iosa et al., 2021; Jehkonen et al., 2000; Kalra et al., 1997). Found in about one quarter to one third of all patients with acute right hemisphere stroke (Becker & Karnath, 2007; Rengachary et al., 2011), spatial neglect poses a great challenge to our health system. About one third of these acute neglect patients manifest chronic neglect more than a year after the neurological incident (Karnath et al., 2011). One of the most pressing questions after a stroke is whether an individual patient will recover in the long term. An early differentiation between patients who will vs. who will not (fully) recover from spatial neglect is therefore of great relevance for the affected patients, their relatives, as well as clinicians. Improved prognosis could not only provide a more realistic expectation of one’s own disease course but would also help guide individually tailored treatment.

Previous research evaluating potential predictors for neglect recovery had suggested initial neglect severity (Moore et al., 2021; Samuelsson et al., 1997; Stone et al., 1992), lesion size (Hier et al., 1983; Jehkonen et al., 2007; Levine et al., 1986), and lesion location (Farnè et al., 2004; Hier et al., 1983; Samuelsson et al., 1997). In addition, demographic data such as age and sex are commonly investigated in prediction studies of post-stroke recovery (Hier et al., 1983; Hope et al., 2013). However, the different studies provided heterogeneous and even contradictory results with respect to the most predictive factors for neglect recovery. Studies are missing that employ modern machine learning-based methods while using behavioral, demographic, and anatomical lesion data to predict persistent neglect. In the present longitudinal study, we thus investigated the clinically important question whether behavioral, demographic, and structural stroke lesion information acquired during the acute phase of stroke can predict long-term neglect prognosis. We examined the predictive values of imaging and non-imaging data in models of different variable combinations. Beyond, in a previous investigation of our group (Karnath et al., 2011), it was examined whether acute anatomical scans could predict the recovery of spatial neglect 1.4 years post-stroke by using voxel-wise lesion analysis approaches (Rorden et al., 2007; Rorden & Karnath, 2004). At the cortical level anterior parts of the superior and middle temporal gyri and subcortically the basal ganglia were found to be critically involved when neglect behavior became a chronic disorder (cf. Fig. 2C in Karnath et al., 2011). Therefore, lesion to these structures was also investigated as a possible predictor for neglect prognosis.

## Methods

### Patient sample

Neurological patients consecutively admitted to the Center of Neurology at Tübingen University were screened for an acute right hemispheric stroke. Patients with a left hemispheric stroke, with diffuse or bilateral brain lesions, with lesions restricted to the brainstem or cerebellum, with tumors, with no visible demarcations, or without acute imaging data were not enrolled. We included 72 patients in total. All patients were screened for spatial neglect on average 5.8 days (SD 6.4 days) post-stroke the first time. Patients suffering from spatial neglect were re-investigated in the chronic phase of stroke approximately 1.6 years (566.5 days ± SD 398.3 days), but not before 6 months, after the initial examination. According to the diagnostic tests of both acute and chronic time points (cf. next paragraph), patients were assigned to three different groups: the “*chronic*” group consisted of 12 patients who showed spatial neglect during both acute and chronic phases; the “*recovered*” group consisted of 30 patients who showed spatial neglect during the acute but no longer during the chronic phase of stroke; the “*control*” group consisted of 30 stroke patients who showed no neglect symptoms in the acute phase of stroke. Twenty-five patients (5 chronic and 20 control patients) of these 72 patients were also included in the creation of the chronic neglect ROI map in the preceding study of our group (Karnath et al., 2011). All patients did consent to the participation in the study, which was approved by the Ethic Commission of the Medical Faculty of the University of Tübingen and was performed in accordance with the revised Declaration of Helsinki of 1964. Demographic and clinical data of all patients are presented in Table 1.

**Table 1.**
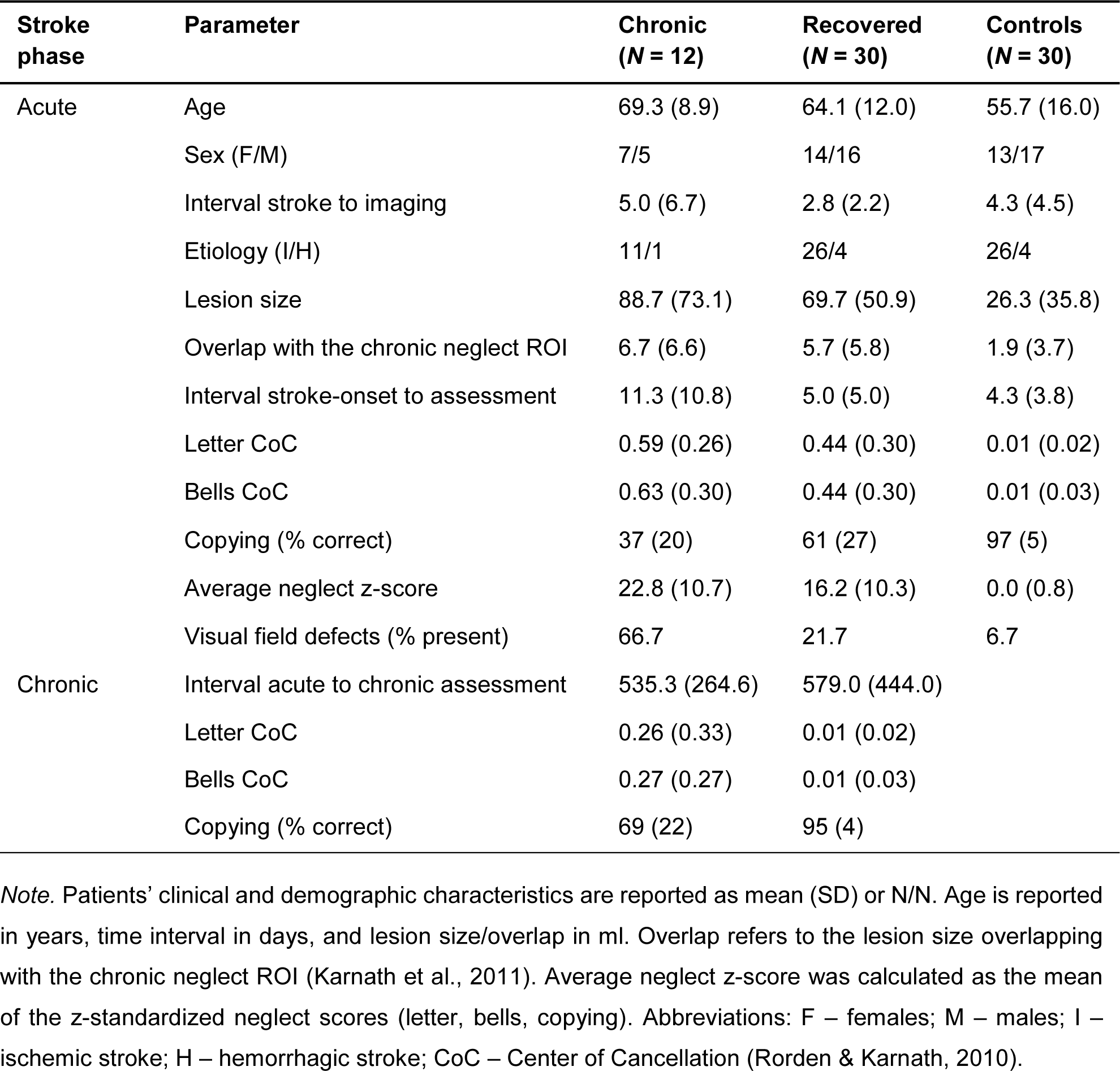
Sample characteristics.

### Behavioral data

Visual field defects were examined by the common neurological confrontation technique. Patients were assessed using the following clinical tests to quantify the severity of spatial neglect: letter cancellation task (Weintraub & Mesulam, 1985), bells cancellation test (Gauthier et al., 1989), and the copying task (Johannsen & Karnath, 2004). All three tests were presented on a horizontally oriented A4 sheet of paper. For both cancellation tests, we calculated the “center of cancellation” (CoC; Rorden & Karnath, 2010). The CoC score measures neglect severity in a continuous manner, while capturing both the number and location of omissions. CoC scores greater than 0.081 (letter) or 0.083 (bells) were considered as being pathological for a left-sided neglect (cf. Rorden & Karnath, 2010). In the copying task, omission of at least one of the contralateral features of each figure was scored as 1, and omission of each whole figure was scored as 2; one additional point was given when contralesional figures were drawn on the ipsilesional side of the test sheet; the maximum score was 8. A score of minimum 2 (i.e. ≥ 25% omissions) indicated neglect behavior in the copying task (Johannsen & Karnath, 2004). A patient was considered of showing spatial neglect when at least one out of the three diagnostic tests was pathological. Following this reasoning, a patient was only considered as “control” if all three diagnostic tests were available and non-pathological.

### Imaging data

Brain lesions were demonstrated by clinical MRI (68%) or CT (32%) scans that were acquired for diagnostic reasons at admission. For patients who underwent MRI scanning, we used diffusion-weighted imaging (DWI) within the first 48h post-stroke and a T2-FLAIR sequence when imaging was conducted 48h or later after stroke onset. For image preprocessing and data analyses, we used MATLAB R2019a and R2023a (The MathWorks, Inc., Natick, USA). Stroke lesion delineation was performed via the semi-automated “Clusterize Toolbox” (Clas et al., 2012; de Haan et al., 2015) for SPM12 (Statistical Parametric Mapping; Wellcome Department of Imaging Neuroscience, London, UK). The resulting binary lesion map was transformed into standard MNI space (Montreal Neurological Institute) with voxels of size 1×1×1mm using the “Clinical Toolbox” (Rorden et al., 2012) by normalizing the anatomical brain scan together with the lesion map to age-matched templates. In thirteen cases, only manual delineated lesion maps created for previous studies (by using MRIcron software; https://www.nitrc.org/projects/mricron) were available in MNI space. Finally, all lesion maps were restricted to cerebrum tissue, i.e. ventricles, cerebellum etc. were masked. A simple overlap of the brain lesions is presented in Figure 1.

**Figure 1.**
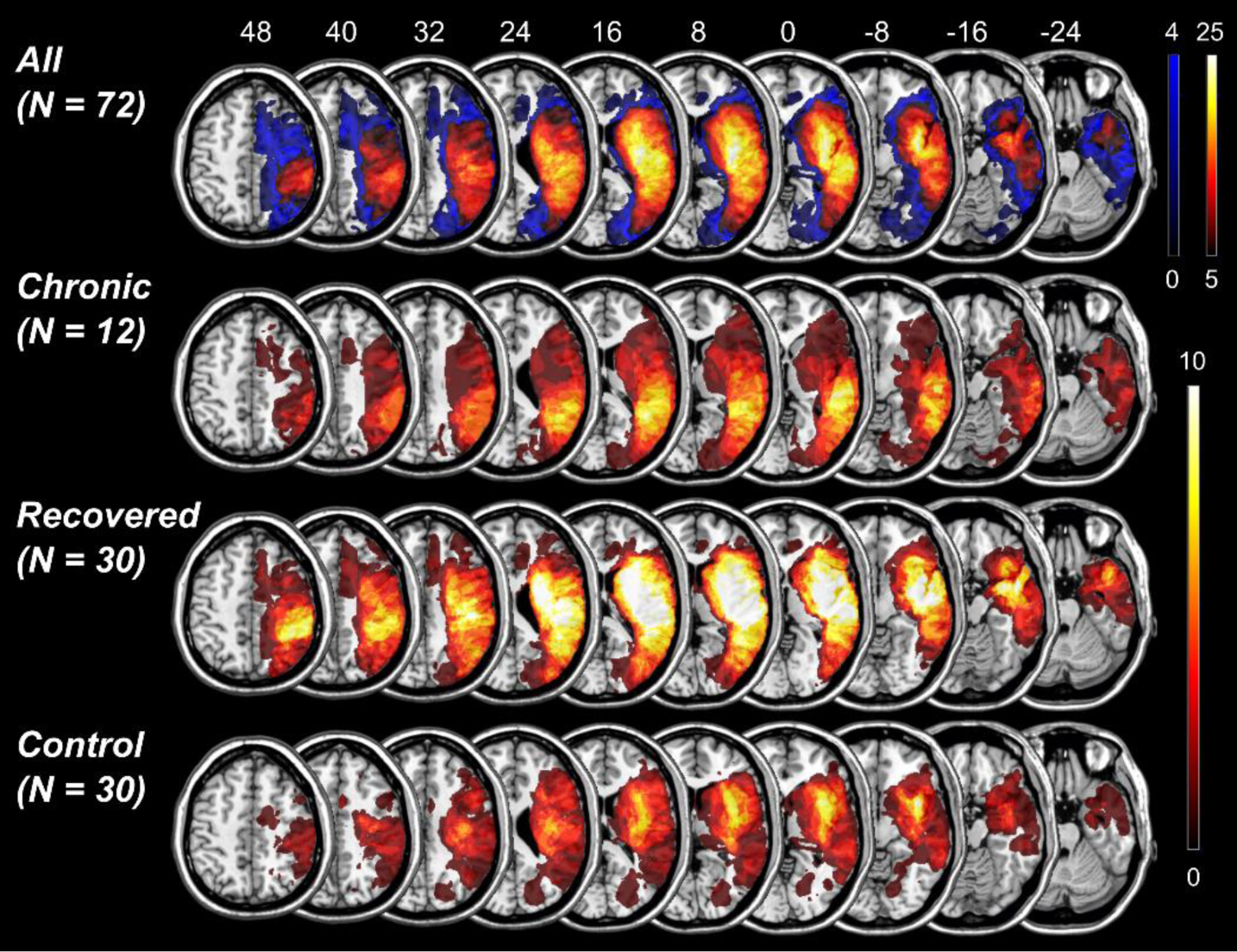
Lesion overlay. Simple overlay maps are presented for the whole sample (upper row), chronic neglect patients only (second row), recovered neglect patients only (third row), and control patients only (lowest row) on axial slices of the ch2-template using MRIcron software. The color legend represents the number of patients with damage to a voxel. For the total sample, bluish colors refer to voxels that were damaged less than five times among all patients and therefore were excluded from further analyses. Numbers above the brain slices refer to z-coordinates in mm in standard MNI space.

### Data analysis

#### Target variables

We calculated different scores to quantify the prognosis of spatial neglect and investigated which one can most accurately be predicted by our algorithm and predictors. We tested the following three variables that describe different aspects of neglect in the chronic phase of stroke: chronic z-score, z-score difference, and effectiveness of recovery (Fig. 2A).

**Figure 2.**
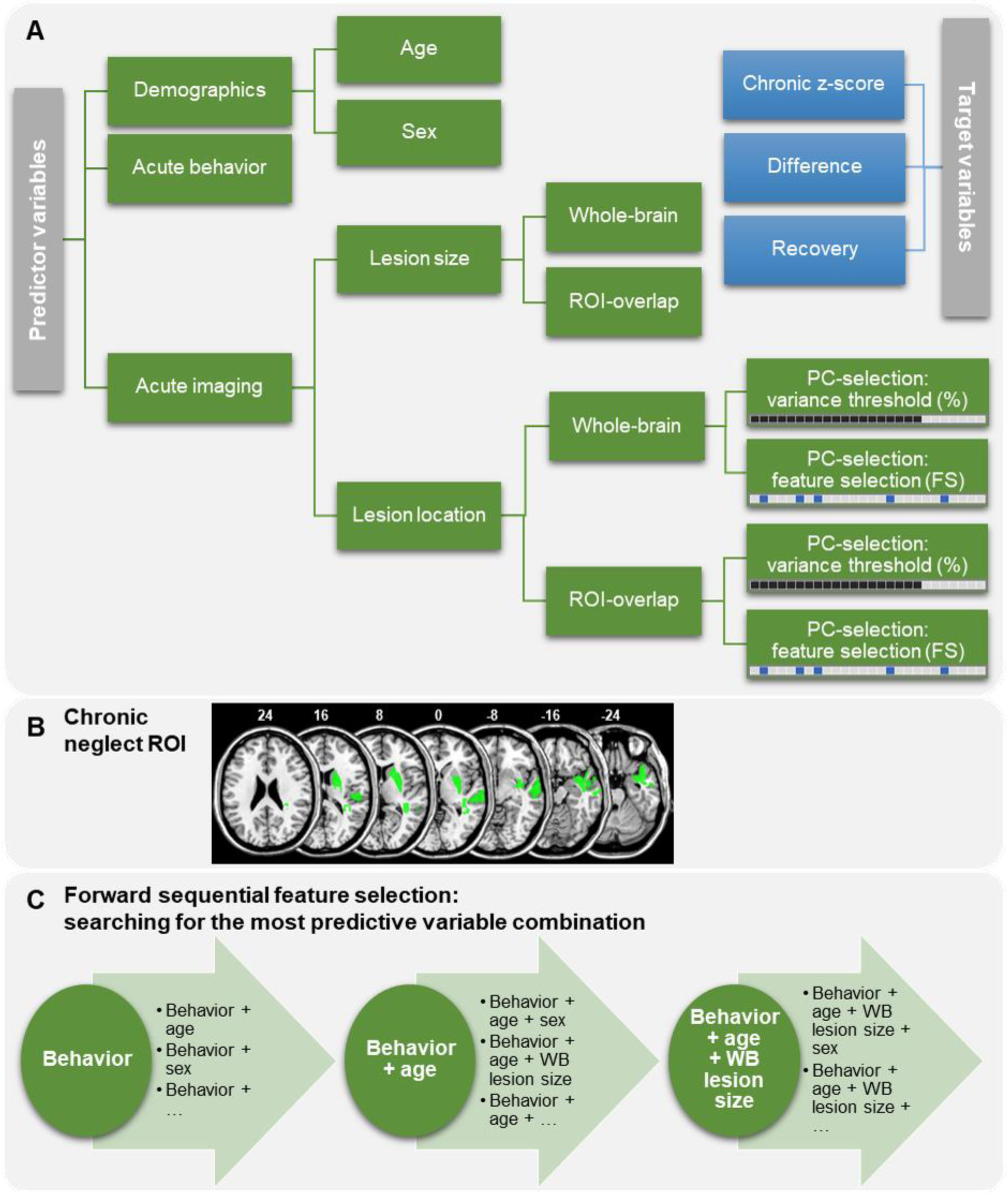
Variables and model selection procedure. **(A)** Predictor variables (green) and target variables (blue) are presented. Principal components of the lesion location data were selected in two different ways: we either kept all PCs that cumulatively explained a certain proportion of the total imaging variance (black squares, in the following identifiable by “%”), or we identified the five most important PCs, i.e. that were most strongly associated with the target variable, filtered by a feature selection approach (blue squares, in the following identifiable by “FS”). **(B)** The chronic neglect region of interest (cf. Fig. 2C in Karnath et al., 2011) is depicted in green in MNI space (numbers refer to the z-coordinates in mm). The overlap of an individual lesion map with the chronic neglect ROI was used to derive the ROI-based predictor variables. **(C)** The forward sequential feature selection is exemplarily illustrated; the first iteration represents the selection of the most predictive single variable (in the presented example: acute behavior), followed by multiple iterations of variable combinations. As long as the prediction error could further be minimized, the number of predictors was increased. The model selection procedure was designed to minimize the prediction error of chronic and recovered neglect patients (excluding control patients). For each target variable, this model selection procedure was applied individually.

#### Chronic z-score

To describe the overall neglect severity with only one score, we calculated the z-scores for the letter and bells cancellation tasks as well as for the copying task by using the acute data of the control patients for standardization. Neglect severity was then determined as a continuous variable by averaging the three z-transformed test scores, separately for the initial and chronic values. The chronic z-score served as one target variable and represents the severity of persistent neglect itself.

#### Difference between acute and chronic z-scores

The acute z-score was calculated equivalent as described for the chronic z-score. The z-score difference was then calculated by subtracting the chronic z-score from the acute z-score. This target variable thus describes the quantitative difference, which is the direct improvement, in neglect severity between the acute and chronic stages.

#### Effectiveness of recovery

To evaluate the recovery rate of spatial neglect between initial and chronic post-stroke phases, we calculated the effectiveness of recovery. Calculation was based on a previously reported formula (adapted from Grasso et al., 2005; Shah et al., 1990; for details, see Supplementary Material). In short, this variable depicts the proportion of potential recovery that was actually achieved, dependent on the acute neglect severity. Effectiveness of recovery was 100% if a patient’s neglect had entirely resolved in the chronic phase; recovery was 0% if a patient’s neglect severity score did not improve at all. The recovery score was calculated for each diagnostic test (letter, bells, and copying) separately and was averaged afterwards.

Because control patients are very unlikely to exhibit neglect behavior during the chronic phase of stroke, we set their chronic neglect z-scores to zero and their recovery scores to 100%.

#### Predictor variables

We tested the predictive value of the severity of acute neglect behavior, demographic data, volume of the brain injury, and its location (Fig. 2A). To operationalize acute neglect behavior, we used the mean acute CoC of letter and bells cancellation tasks as this measure can be easily obtained in a clinical context. For the demographic data, we used age at stroke-onset and sex. Further, we tested the whole-brain lesion size (“*WB lesion size*”). To determine the validity of a previously suggested anatomical map as a predictor of chronic neglect and to compare it to the predictive value of the total stroke lesion, we further created lesion maps that included only those portions of the lesion that overlapped with this map, hereafter labeled as “chronic neglect region of interest (ROI)”. The latter comprised at the cortical level parts of the superior and middle temporal gyri and subcortically the basal ganglia (especially the putamen) which had been reported to be critically involved when neglect behavior became a chronic disorder (Fig. 2B above; Karnath et al., 2011). Besides the whole-brain lesion size, we therefore also investigated the size of the individual lesion overlaps with the chronic neglect ROI (“*ROI lesion size*”) as a measure of ROI-based lesion extent. This feature-can also be seen as the relative damage of the total chronic neglect ROI.

Furthermore, we tested different variables of lesion location to investigate the impact of lesion topography on prediction accuracy. Voxels damaged less than 5 times among all patients were excluded to ignore rarely affected voxels with low statistical power. We ran a principal component analysis (PCA) to reduce the dimensionality of each map variant (i.e. 1. whole-brain lesion maps and 2. ROI-based overlaps). Principal components (PCs) were previously found to yield more accurate predictions compared to voxel-wise maps (Kasties et al., 2021). We implemented two separate approaches of component selection to keep the target variable-specific most predictive PCs. Firstly, we selected PCs that cumulatively explained at least a specified amount of total variance (see Fig. 2A). For each target variable and map variant separately, we tried different thresholds and used the winning threshold – that produced the most accurate predictions – for further analyses (for details, see Supplementary Material). These lesion location variables are termed “*WB-%-PCs*” and “*ROI-%-PCs*” in the following. Secondly, we selected PCs that were identified as being most relevant for the target variable (see Fig. 2A). Again, for each target variable and map variant separately, we tried three different feature selection filter methods (for details, see Supplementary Material). We then selected the five most important components that were most strongly associated with the target variable, identified by the winning filter method. The resulting lesion location variables are termed “*WB-FS-PCs*” and “*ROI-FS-PCs*” in the following. One of our questions was whether the ROI-based lesion size and/or map might be more informative than the whole-brain lesion size/map, as it was proposed to contain only regions critical for neglect chronicity.

#### Prediction algorithm

First, each target/predictor variable was set to the range of 0 to 1. To predict neglect prognosis, we applied a supervised learning algorithm, support vector regression (SVR) (Brereton & Lloyd, 2010; Smola & Schölkopf, 2004). We used the nonlinear radial basis function (RBF) kernel, as nonlinear kernels were found to be more accurate than linear ones in previous lesion-behavior studies (Hope et al., 2018; Zhang et al., 2014); in accordance, predictions were overall less accurate using a linear kernel in a pilot investigation of the current study. We implemented an epsilon-SVR using the “libsvm” package for MATLAB (Chang & Lin, 2011). The algorithm is a custom script for MATLAB that is based on the algorithm described and visualized in Röhrig et al. (2022). We used either one or multiple independent predictor variables to predict one dependent target variable. To get generalized results that might not only be applicable to the current sample, we applied a nested cross validation (CV) approach (Krstajic et al., 2014; Varoquaux et al., 2017) with five iterations in the outer loop and four iterations in the inner loop. A grid search for optimizing the hyperparameters C and gamma was implemented within the inner loop (*C* = 2^(−5)^, 2^(−4)^, …, 2^15^; *gamma* = 2^(−15)^, 2^(−14)^, …, 2^5^). In the whole model selection procedure, the aim was to minimize the prediction error, i.e. the mean squared error (*MSE*). The winning model of the inner loop (determined by the lowest *MSE*) was tested on the unknown, hold-out test set of the outer loop. In the end, each patient was predicted once during the outer loop. This procedure was repeated 10 times with different sample randomizations to further generalize the algorithm as the random assignment of patients to the training-, validation-, and test-sets influences the model performance. The out-of-sample predictions obtained by these model repetitions were averaged for each patient (i.e. model averaging; Arlot & Celisse, 2010; Varoquaux et al., 2017). These final predictions were used to calculate the overall model fit. We again used the *MSE* to select the best model and report the goodness of fit (coefficient of determination, *R²*) and the Pearson correlation coefficient (*r*) as additional measures of model performance; the *R²* gives the proportion of total variance explained by a model, whereas *r* describes the linear relationship between actual and predicted test scores. We did this whole procedure for each target variable and predictor set separately.

Previous studies predicting the recovery of cognitive deficits after stroke represent an inconsistency in how to deal with control patients, that is, whether to include (Hope et al., 2013; Lunven et al., 2015; Umarova et al., 2016) or not to include (Halai et al., 2020; Hillis et al., 2018; Stone et al., 1992) patients without a behavior/deficit of interest. On the one hand, patients that do not elicit a certain deficit can help the algorithm to learn which features are not essential for the deficit (cf. Hope et al., 2013). On the other hand, we aimed to predict chronic neglect, which means that for control patients, the chronic outcome cannot be pathological in the chronic phase since it was already non-pathological in the acute phase of stroke (in addition to the circumstance that we estimated the exact chronic scores for control cases). For any algorithm, prediction of controls’ chronic severity would therefore be quite easy (especially if acute behavior is a predictor). The very good predictions for the control subsample would thus artificially enhance the overall model accuracy and give an overoptimistic image. For this reason, we chose a combination of both variants: we used the total sample including control patients to train the algorithm, but we predicted only non-control patients (chronic and recovered) – both in the inner and outer loop of the nested CV. Hence, the model selection approach was built to minimize the prediction error for neglect patients only. This way, we could answer which variables help to distinguish chronic versus recovered patients and not neglect versus non-neglect patients (as it might be relevant in the prediction of acute neglect severity, cf. Röhrig et al., 2022). To this end, the calculation of model performance metrics (mean *MSE*, *R²*, *r*) and hence the whole model selection process (see below) was solely based on predictions of neglect patients (without control patients).

#### Model selection procedure

To test which predictors can most accurately predict neglect prognosis, we evaluated different model variants. For each target variable separately, we investigated the predictive values of (i) single variables, (ii) combinations of thereof, and (iii) full models. Models were selected according to the smallest *MSE* (and thus to the largest *R²*). For the first approach, we tested each predictor individually. For the second approach, we implemented a forward sequential feature selection method that searched for the most predictive variable combination (Fig. 2C). Starting with the predictor that achieved the smallest *MSE* among all single variables, we added a second variable to that first selected predictor. In this feature selection iteration, we added each remaining variable to the first selected predictor once. Among these models, we selected the variable that could further (or most strongly) minimize the *MSE* as the second selected predictor. The procedure of increasing the number of predictors was repeated as long as the *MSE* could further be minimized. For the third approach, we investigated full models, i.e. including all predictor variables at once. Because different lesion location variables (*WB-%-PCs*, *ROI-%-PCs*, *WB-FS-PCs*, *ROI-FS-PCs*) cover overlapping information, we included at most one of them in each model.

## Results

### Clinical and demographic variables

As expected, we detected higher neglect severity (i.e. a higher chronic z-score) in the chronic stroke phase for patients of the chronic subsample and no neglect symptoms for recovered patients (cf. Tab. 2). In line with that, findings revealed a higher recovery rate (i.e. a higher effectiveness of recovery) for recovered than for chronic patients, whereas the improvement from the acute to the chronic stroke phase (i.e. z-score difference) did not statistically differ between recovered and chronic patients. Furthermore, compared to recovered neglect patients, chronic patients were on average older (*t* = 1.37), more often female (*chi²* = 0.47), had larger lesions (*t_unequal_* = 0.82), and larger lesion overlaps with the chronic neglect ROI (*t* = 0.45); however, these effects were not statistically significant (*p* > 0.05; cf. Tab. 1). In contrast, chronic patients had significantly more often visual field defects than recovered patients (*chi²* = 5.79, *p* = 0.016; cf. Tab. 1).

**Table 2.** Target variables of neglect prognosis.

| Parameter | Chronic<br>(*N* = 12) | Recovered<br>(*N* = 30) | *t* | *p* |
| --- | --- | --- | --- | --- |
| Average chronic neglect z-score | 9.8 (10.4) | 0.0 (0.8) | 3.29* | **0.007** |
| Difference acute - chronic z-scores | 14.0 (9.8) | 16.5 (10.5) | -0.71 | 0.48 |
| Effectiveness of recovery (%) | 59.1 (27.9) | 93.3 (12.5) | -4.08* | **0.001** |

*Note.* Target variables are reported for the chronic and recovered neglect subsamples. The chronic z-score represents the chronic neglect severity itself; the z-score difference equals the direct improvement from the acute to the chronic stage of stroke; and the effectiveness of recovery describes the proportion of potential recovery that was actually achieved. Statistical results were obtained from two-sample two-tailed *t*-tests; asterisks mark unequal variances *t*-tests and bold *p*-values highlight significant results.

### Neglect prognosis

For a better understanding of the following results, please have in mind that an ideal model would result in *MSE* = 0, *R²* = 1, and *r* = 1. Note that *R²* usually ranges between 0 and 1, but as we predicted unseen data during the nested CV, *R²* can be negative in case of less accurate predictions compared to when always predicting the average test score. A negative *R²* and a negative *r*, however, are of no interest as only positive scores represent optimal predictions.

All prediction accuracies are reported in detail in Tables S1, S2, and S3 in the Supplement. Overall, we found that full models did not outperform the best single predictor models. However, the feature selection process did further improve the prediction accuracy by detecting a predictive variable combination (but only for the z-score difference).

#### Chronic z-score

The single variable most predictive for the chronic z-score was *WB-FS-PCs* (*MSE* = 0.022 ± SD 0.057, *R²* = 0.55, *r* = 0.77 with *p* < 0.001; Fig. 3A), which is the lesion location described by the five most important PCs derived from the whole-brain lesion maps. Besides this variable, the single predictors *ROI lesion size* (*R²* = 0.25) and *ROI-FS-PCs* (*R²* = 0.07), i.e. the lesion location described by the five most important PCs derived from the individual ROI overlaps, did also explain some proportion of the total variance, representing predictive power. Nevertheless, the prediction accuracy could not further be improved during feature selection (see Fig. 3B). The most accurate full model also included the variable *WB-FS-PCs* (*MSE* = 0.047 ± SD 0.124, *R²* = 0.06, *r* = 0.26 with *p* = 0.09; Fig. 3A).

**Figure 3.**
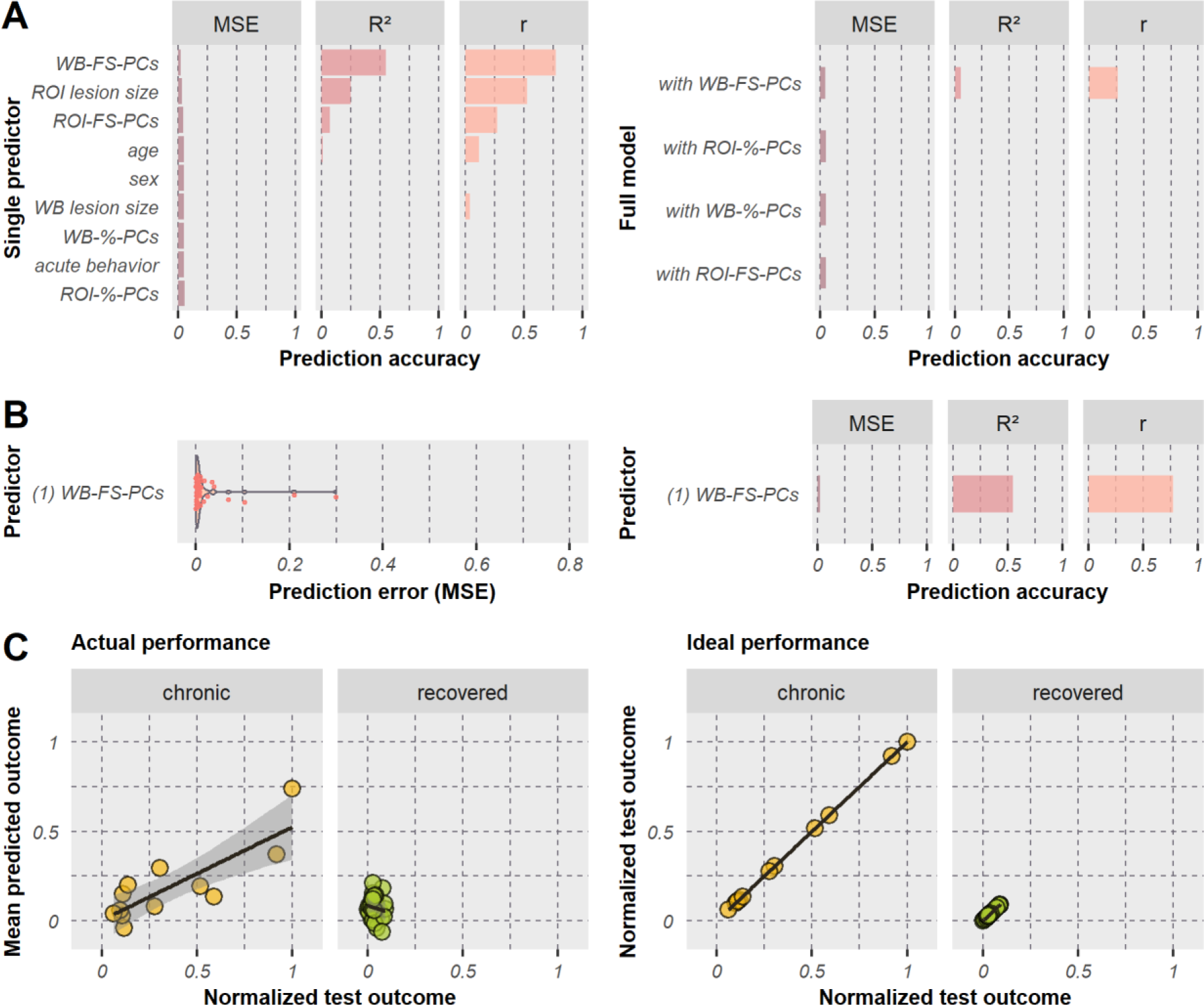
Model performances when predicting the chronic z-score. **(A)** Results are illustrated for single predictors (left) and full models (right); full models included all single variables but only one lesion location variable as predictors. Lesion location variables consisted of principle components (*PCs*) derived from the whole-brain (*WB*) or ROI-based lesion maps, whereby the PCs were either selected according to a specific proportion of cumulatively explained variance (*%*) or the five most important PCs were filtered by a feature selection approach (*FS*). Models are sorted by increasing prediction error for chronic and recovered neglect patients (*N* = 42). Bar plots represent the mean squared error (*MSE*), coefficient of determination (*R²*), and Pearson correlation coefficient (*r*). Note that only positive values are depicted since a negative *R²* is not informative except for an insufficient model performance and a negative *r* represents the opposite direction of interest. **(B)** Results are shown for the predictor selected by feature selection. The violin plot (left) illustrates the distribution of the individual prediction errors. **(C)** Test scores versus predicted scores are shown for the best performing model (*WB-FS-PCs*). Scatter plots illustrate either actual obtained out-of-sample predictions (left) or perfect predictions (in case of 100% accuracy; right), each for chronic (*N* = 12) and recovered (*N* = 30) neglect patients separately.

This leaves the most predictive single variable as the best model, which explained 55% of the total variance of neglect patients. This model revealed predictions with a strong relationship to the test scores for chronic patients (*r* = 0.82, *p* < 0.001; Fig. 3C). Although the correlation for recovered patients was not powerful (*r* = −0.14, *p* = 0.47), predictions clustered within the ideal range (see Fig. 3C). The five predictive components (43.1ml, 6.4% cumulative explained imaging variance) involved gray matter regions (Brainnetome Atlas; Fan et al., 2016) mainly in the middle and superior temporal gyri (MTG/STG), insular gyrus, basal ganglia (putamen, globus pallidus, dorsal caudate), inferior parietal lobule (IPL), and postcentral gyrus (Fig. 4A). With respect to white matter fiber tracts (JHU ICBM atlas; Mori et al., 2008), mainly the external and internal capsules, superior and posterior corona radiate, and superior longitudinal fasciculus (SLF) were involved. Further details are reported in Table S4 in the Supplement. Findings revealed that lesion location quantified by only five PCs derived from the whole-brain lesion map is the strongest predictor for chronic neglect severity, indicating that the patient’s lesion topography is more important than other clinical or demographic information.

**Figure 4.**
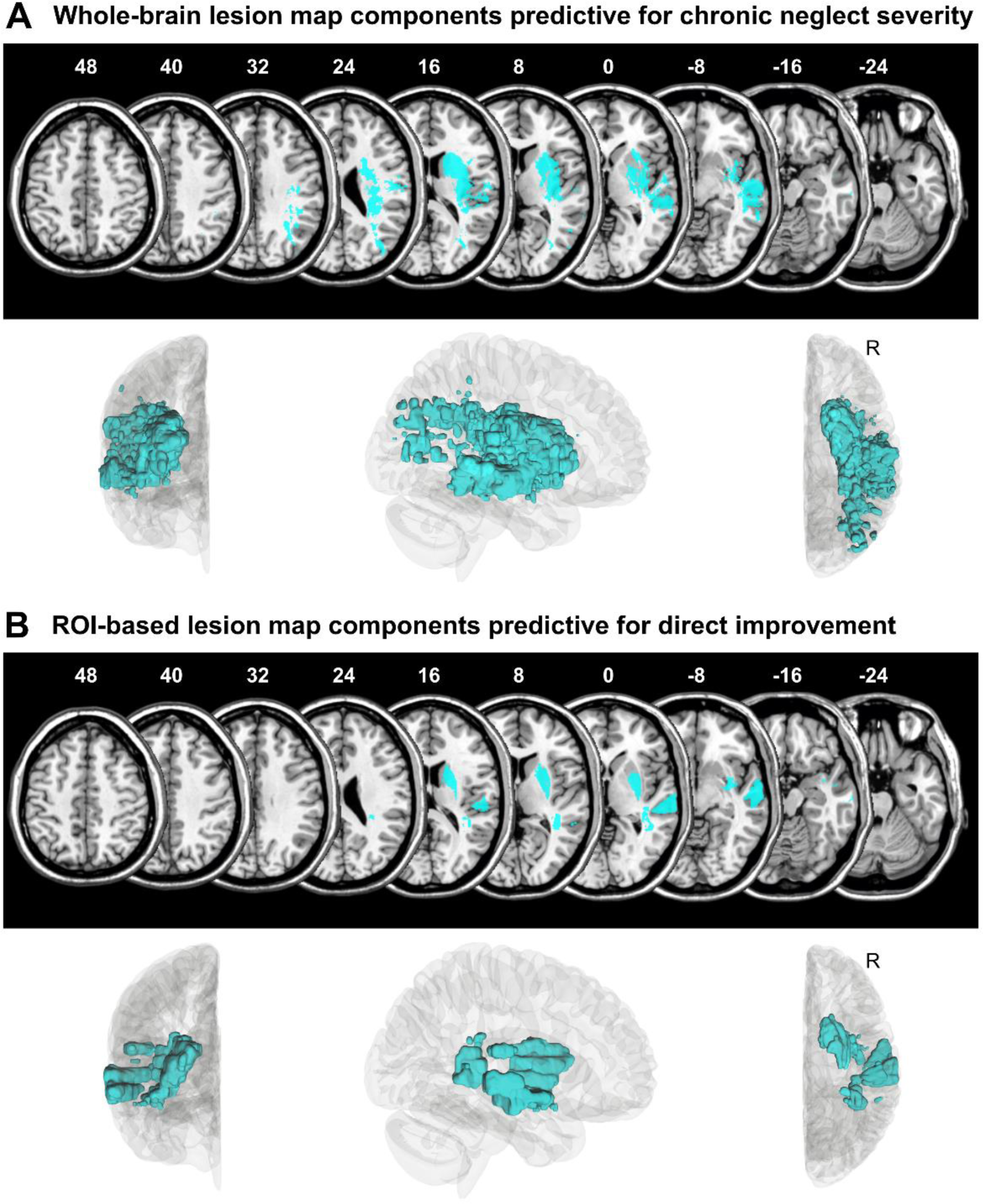
Predictive lesion locations. The five principal components (PCs) are visualized together (cyan) that were obtained from (**A**) the whole-brain lesion map or (**B**) the lesion overlap with the chronic neglect ROI, that were selected based on the respective target variable by a feature selection filter method, and that were found to be together predictive for (**A**) the chronic z-score respectively (**B**) the z-score difference. Upper row each: PCs are presented on axial slices in standard MNI space on the ch2-template via MRIcron software, with z-coordinates above each slice. Lower row each: PCs are shown within the right hemisphere (“*R*”) of a 3D glass brain in anterior, right, and superior views via DSI studio software.

#### Difference of acute z-score minus chronic z-score

The z-score difference, i.e. the direct improvement of neglect severity between acute and chronic phase of stroke, was best predicted by the variable *acute behavior*, which is the initial neglect severity (*MSE* = 0.026 ± SD 0.06, *R²* = 0.59, *r* = 0.77 with *p* < 0.001; Fig. 5A). In contrast to the chronic z-score, the feature selection detected a combination of three variables as the most predictive variable combination for the z-score difference. In addition to the *acute behavior*, selected variables were *ROI-FS-PCs*, i.e. the lesion location described by the five most important PCs derived from the individual lesion overlaps with the chronic neglect ROI, and *ROI lesion size*, i.e. the size of the stroke lesion overlapping with the chronic neglect ROI (*MSE* = 0.022 ± SD 0.048, *R²* = 0.66, *r* = 0.81 with *p* < 0.001; Fig. 5B). Although all full models performed similarly well, the most accurate full model included variable *ROI-FS-PCs* (*MSE* = 0.028 ± SD 0.071, *R²* = 0.56, *r* = 0.75 with *p* < 0.001; Fig. 5A).

**Figure 5.**
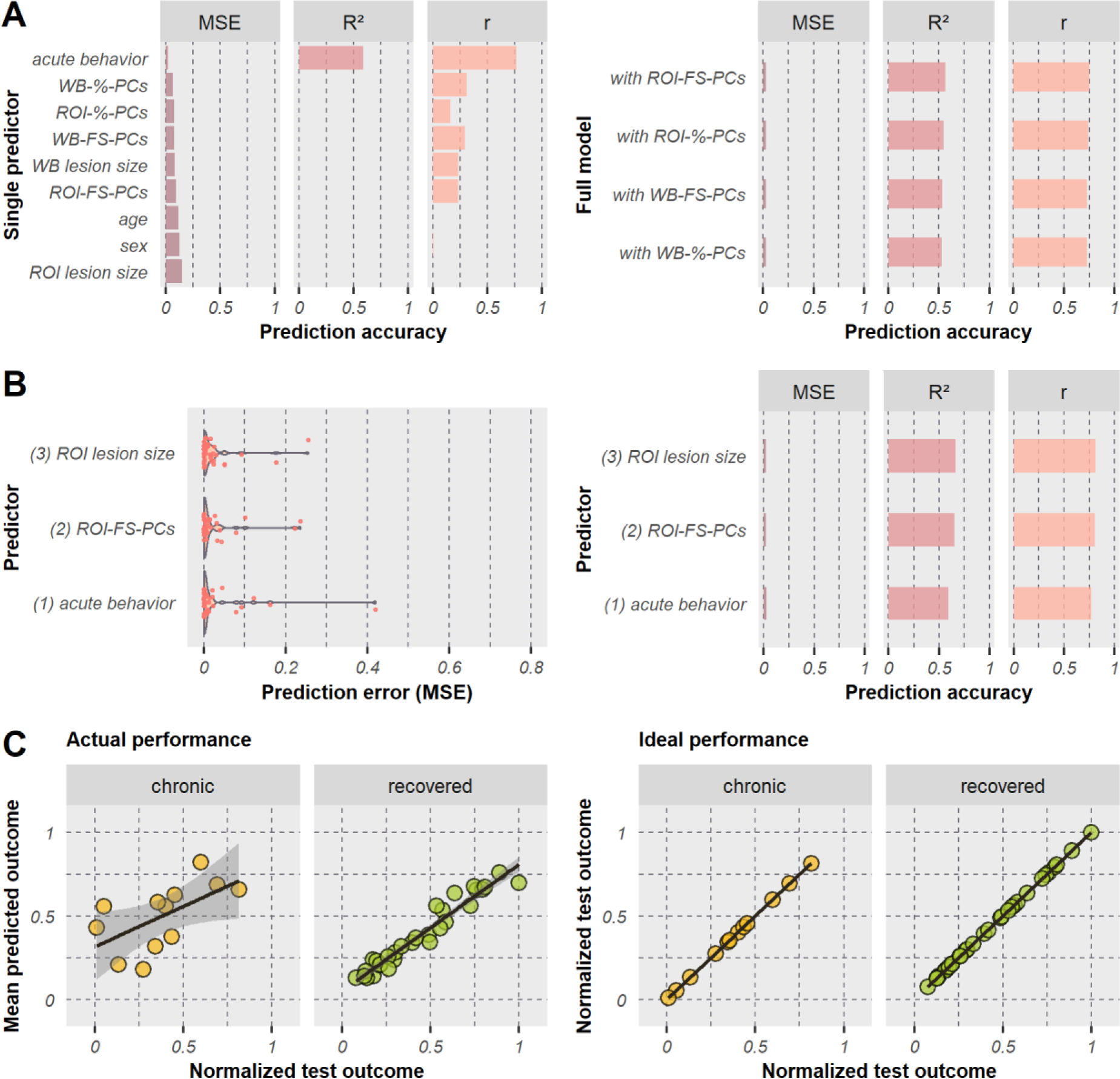
Model performances when predicting the z-score difference. **(A)** Results are illustrated for single predictors (left) and full models (right), and **(B)** for predictors selected by feature selection; the number in front of the variable name represents the iteration of the forward sequential feature selection in which the variable was chosen. **(C)** Test scores versus predicted scores are shown for the best performing model (*acute behavior, ROI-FS-PCs, ROI lesion size*). For all further details, see legend of Figure 3 above.

Overall, the best model for predicting the z-score difference included the following predictors: *acute behavior*, *ROI-FS-PCs*, and *ROI lesion size*. This model explained 66% of the total variance of neglect patients, leaving it as the overall best model among all target variables. Looking at the model fit separated by group, chronic as well as recovered patients received good predictions, yielding correlations between actual and predicted test scores of *r* = 0.60 (*p* = 0.041) for chronic and *r* = 0.97 (*p* < 0.001) for recovered patients (Fig. 5C). The five predictive components (15.5ml, 19.2% cumulative explained imaging variance) mainly involved gray matter regions in the MTG/STG, basal ganglia (putamen, globus pallidus, dorsal caudate), IPL, and insular gyrus as well as white matter fiber tracts as the external and internal capsules, posterior thalamic radiation, and SLF (Fig. 4B; further details in Tab. S4). The findings highlight that regions covered by the chronic neglect ROI were more predictive than the whole-brain stroke lesion when predicting the z-score difference. Location and size of lesion overlaps with the chronic neglect ROI added relevant information to the acute neglect behavior, although they were not predictive among the single predictors. In the end, the initial neglect severity was found to be the best prognostic factor.

Some patients of the current investigation were also included in the creation of the chronic neglect ROI map of the previous study (Karnath et al., 2011; see above section ”Patient sample”). Since ROI-based predictor variables were found to be within the best performing model, we addressed possible concerns due to the sample overlap by additionally calculating the final measures of model performance without predictions of the overlapping chronic patients (*N* = 5). We found no decrease in model performance (*MSE* = 0.014 ± SD 0.032, *R²* = 0.79, *r* = 0.91 with *p* < 0.001; Fig. S1 in the Supplement), indicating that the overlapping patients did not boost the performance. However, note that the improvement of accuracy, which was even achieved, was due to the reduction of the chronic subsample, as chronic patients were less predictable than recovered ones.

To investigate whether every selected predictor contributes significantly to the overall model performance, we implemented permutation tests with 5000 iterations each. Acute neglect behavior (*p* < 0.001) and ROI-based lesion location (*p* = 0.047) were found to be individually important for the prediction accuracy, whereas size of the ROI-based lesion overlap did not achieve significance (*p* = 0.061).

#### Effectiveness of recovery

Prediction errors were overall larger for models predicting the effectiveness of recovery, indicating that the recovery score can be predicted least well among the evaluated target variables. The variable *WB lesion size*, i.e. the volume of the whole-brain stroke-induced damage, resulted in the lowest prediction error among all single predictors (*MSE* = 0.066 ± SD 0.163, *R²* = −0.06, *r* = 0.10 with *p* = 0.55; Fig. 6A). As for the chronic z-score, the feature selection for the recovery score detected no variable combination that outperformed the most predictive single predictor (Fig. 6B). The model with the lesion location variable *ROI-%-PCs* served as the best performing full model (*MSE* = 0.068 ± SD 0.153, *R²* = −0.10, *r* = −0.31 with *p* = 0.045; Fig. 6A).

**Figure 6.**
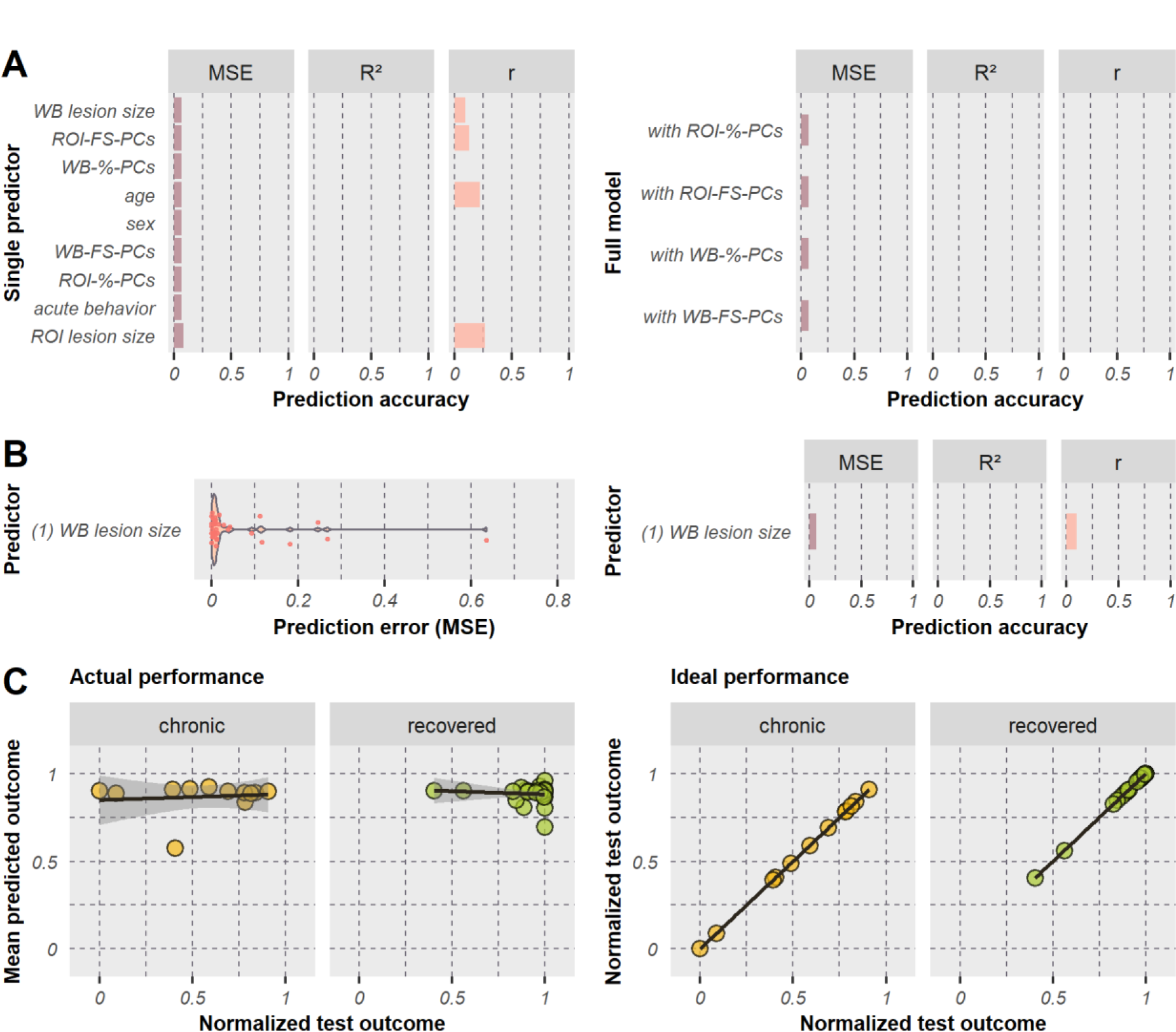
Model performances when predicting the effectiveness of recovery. **(A)** Results are illustrated for single predictors (left) and full models (right), and **(B)** for the predictor selected by feature selection. **(C)** Test scores versus predicted scores are shown for the best performing model (*WB lesion size*). For all further details, see legend of Figure 3 above.

Although the model including the predictor *WB lesion size* yielded the lowest error for predicting effectiveness of recovery, the overall performance was not sufficient to be meaningful. This is illustrated by the poor model fit for chronic (*r* = 0.11, *p* = 0.74) and recovered patients (*r* = −0.11, *p* = 0.58; Fig. 6C). In fact, findings revealed that none of the models that aimed to predict the effectiveness of recovery was able to explain some proportion of total variance (no positive-valued *R²*). To summarize, none of the models predicting neglect recovery was detected to achieve meaningful predictions, which also weakens the potential prognostic value of the whole-brain lesion size.

## Discussion

In the present study, individual long-term outcome of spatial neglect was predicted based on acute patient data in a sample of chronic and recovered patients with neglect, by implementing a repeated nested cross-validation design with feature selection. Out-of-sample predictions suggest that the direct improvement of neglect severity between acute and chronic phases of stroke can be best predicted, followed by chronic neglect severity itself. On the other hand, the effectiveness of recovery turned out to be not sufficiently predictable by the variables and algorithm tested. Among different types of predictors, demographic information (age, sex) seemed to be the least useful. In contrast, initial neglect severity was helpful when predicting the direct improvement of neglect behavior; this variable alone already reached a small prediction error.

Present findings also highlight the prognostic value of imaging data for post-stroke recovery of spatial neglect, which is in line with previous outcomes (Kamakura et al., 2017; Karnath et al., 2011; Lunven et al., 2015; Umarova et al., 2016; for review see Imura et al., 2022). Although lesion size was predictive to a certain extent, findings suggest a superiority of lesion topography. The latter served as a reliable predictor, including brain regions as the middle and superior temporal gyri, insular gyrus, inferior parietal lobule, basal ganglia, and white matter tracts as external capsule, internal capsule, and superior longitudinal fasciculus. However, prediction analyses revealed that the best variable combination comprised the individual neglect severity in the acute phase of the stroke, and size and location of individual lesion overlaps with the previously proposed chronic neglect ROI (Karnath et al., 2011). Using these variables for predicting the direct improvement from the acute to the chronic stroke phase, 66% of the total variance of chronic and recovered neglect patients were explained, making them very promising features in the prediction of individual outcome prognoses. On the other hand, chronic neglect severity could be predicted by using acute lesion data only, reaching an accuracy of 55% explained variance. This is of particularly clinical relevance as patients with severe post-stroke cognitive impairments might be unable to accomplish multiple paper-and-pencil tests to determine the severity of spatial neglect. In such cases, acute lesion information can be used independently from diagnostic behavioral tests to predict chronic outcome already in the acute phase of stroke and eventually guide treatment programs.

Overall, our maximal model performance (i.e. *R²* = 0.66) was remarkably superior to a reported performance regarding visuospatial abilities of *R²* = 0.18, which was obtained by the Disconnectome Symptom Discoverer (DSD) model developed in a recent study on predicting post-stroke cognitive recovery using the structural disconnectome (Talozzi et al., 2023). This performance was achieved for the DSD when predicting individual performances on the Bells cancellation test using a sophisticated model validation approach, including training and validation on different external cohorts. However, as model performance within the training phase did also not exceed 30% explained variance (Talozzi et al., 2023), we can speculate whether structural disconnection data might be less suited for long-term prediction in comparison to lesion anatomy. Contrary, Talozzi and colleagues (2023) observed the opposite, as their structural disconnection model outperformed lesion-based models. In contrast to models investigated by Talozzi and colleagues (2023), our best predictive model included the acute behavior as an important predictor, in addition to lesion-based features. However, it is important to note that the exclusion of acute behavior in the DSD model may only partially contribute to the superior performance of our best model, because another model of the current study achieved a superior model performance of *R²* = 0.55 by using only lesion-based features. Another potential explanation for the superiority of our current approach is that the feature selection procedure implemented – especially the filter method pre-selecting important spatial imaging features – did improve model accuracy. Additionally, the DSD model relies on linear regression, whereas the current approach used support vector regression with a nonlinear kernel. Furthermore, in the samples used for external training and validation of the DSD model, neuropsychological test data (i.e. Bells scores) were obtained in the subacute phase of stroke (within 30-90 days post-stroke), whereas in the current study, chronic behavior (minimum 189 days post-stroke) was predicted using acute imaging. For these reasons, we argue that the fundamental backgrounds of the discussed models are different in nature and allow only limited space for direct comparisons.

### Imaging biomarkers

Simple, one-dimensional imaging information, namely whole-brain (WB) and ROI-based lesion size, was found to be somewhat predictive. The ROI-based lesion size outperformed the WB lesion size and constituted the second-best predictor when predicting the chronic z-score, while, on the other hand, this variable was the least good predictor when predicting the z-score difference or recovery score. During feature selection, the ROI-based lesion size did again outperform the WB lesion size. The reduction of the WB lesion size to chronic neglect regions (Karnath et al., 2011) seems to be beneficial for the accuracy in certain conditions only. Among the best models, variables of lesion size could either not achieve accurate predictions at all (when predicting the recovery score) or did not significantly improve model performance (when predicting the z-score difference). In contrast to a recent investigation on the general predictive power of lesion size and location on stroke severity and outcome (Sperber et al., 2023), we observed that lesion location variables clearly outperformed lesion size variables. On the other hand, when predicting the direct improvement, model performance was only 3.6% less accurate when using only ROI-based lesion size in addition to acute neglect behavior (i.e. without ROI-based lesion location; see Tab. S2 in the Supplement). This indicates that lesion location is only slightly superior to lesion size, which is in accordance with findings by Sperber et al. (2023). However, as size of the ROI-based lesion overlap relies on spatial features of individual stroke lesions (i.e. lesion size is limited to specified brain regions), this feature can further be seen as a one-dimensional lesion location variable. The circumstance that component-based lesion location seems to improve model performances only slightly may be of high interest for clinical settings, as the calculation and implementation of lesion size is more applicable than complex principal components due to the different degrees of dimensionality (cf. Sperber et al., 2023). Hence, a compromise of good accuracy and easy implementation might be lesion size as an overlap of individually damaged voxels with predictive topographical features extracted by our principal component analysis. Corresponding binary anatomical maps are available in the online material.

Regarding complex, high-dimensional lesion location information (i.e. topographic measures), we found that a pre-selection of relevant PCs yielded more accurate predictions compared to PCs that cumulatively explain a specific proportion of total imaging variance. Both the WB lesion map and the ROI-based lesion overlap contained prognostic PCs. We found that the combination of five PCs of the whole-brain lesion map pre-selected by feature selection represents the sole predictor variable of the model most accurately predicting the chronic z-score, i.e. chronic neglect severity. Similarly, for the z-score difference (i.e. the direct improvement), the five PCs of the ROI-based lesion map filtered via feature selection added some predictive value to the so far best-performing model. Results highlight the importance of feature selection and future research on potential imaging biomarkers. Interestingly, both PC selections cover similar brain areas (cf. Fig. 4 above), suggesting that similar regions were predictive for different measures of neglect prognosis. This also strengthens the hypothesis that the described areas might be crucial for persistent symptoms of spatial neglect, supporting findings by Karnath et al. (2011).

The present study revealed that areas covering the MTG/STG, basal ganglia, insula, and IPL were predictive, demonstrating the prognostic value of these gray matter regions and supporting previous findings (Chechlacz et al., 2012; Karnath et al., 2002; Saj et al., 2012). At the connectivity level, we observed that – among other fiber tracts − damage to the SLF was relevant for neglect prognosis, which again is in line with previous observations (Chechlacz et al., 2012; Karnath et al., 2009; Lunven et al., 2015; Thiebaut de Schotten et al., 2014). In fact, damage to these cortical and subcortical structures, in particular, have been reported to be associated with chronic neglect (e.g. Karnath et al., 2011; Lunven et al., 2015; Saj et al., 2012). However, please note that the present study was not designed to determine whether the mentioned structures are frequently lesioned or spared in chronic patients. Instead, the involvement of these structures tells us that their lesion status affects long-term prognosis, that is, either damage or preservation of corresponding voxels is associated with either chronicity or recovery of spatial neglect.

In addition to these anatomical structures which are well known to be related to spatial neglect, we found that lesion status of external and internal capsules was predictive for chronic behavior of neglect patients. In line with this observation, previous research found significantly decreased white matter integrity in right-sided external and internal capsules of recovered (but not chronic) neglect patients compared to non-neglect patients (Lunven et al., 2015), suggesting that damage to these structures might result in acute neglect with high probability of recovery. In contrast, the present analysis did neither find evidence for a major involvement in neglect chronicity for the callosal splenium, as reported by Lunven et al. (2015), nor for the uncinate fasciculus, as reported by Karnath et al. (2011). However, when comparing previous and current results, it should generally be borne in mind that exact comparisons are challenging due to variations in methodologies, research questions, behavioral tests used, time intervals for initial and follow-up assessments and imaging, inclusion or exclusion of control patients, and investigated contrasts (chronic versus recovered, chronic versus control) among studies.

### Challenges of neglect prognosis

Our results clearly showed that the way how chronicity of spatial neglect was calculated (chronic z-score, z-score difference, or effectiveness of recovery) had a major impact on whether the predictor variables had high or low prognostic values. Although direct improvement by simple subtraction of functional score from acute to chronic stage (z-score difference) is a classic measure, it is influenced by floor and ceiling effects (Grasso et al., 2005). Moreover, patients with acute scores representing subtle deficits can potentially improve less than patients with acute scores representing severe deficits (Grasso et al., 2005; Shah et al., 1990). The recovery score addresses these issues by taking the acute neglect severity into account. Nevertheless, the recovery score also has limitations. Patients with 50% recovery might have had a very large direct improvement or almost no direct improvement, depending on the acute behavior. The algorithm built in the current study could not predict such information reliably, possibly because very different patient subsamples were considered as being similar. Another possible explanation why we could not solidly predict the recovery score is that our algorithm would have needed more training examples to reliably detect patterns among the complex recovery scores. Nevertheless, one could try implementing models with multiple target variables in future studies. To put all three variables examined in the present study into context: a patient with severe neglect but minimal recovery achieved the same direct improvement (absolute change in neglect behavior) as a patient with slight neglect symptoms and full recovery. Each test variable was best predicted by different predictor combinations and could be predicted to different degrees, which supports the importance of examining different variables of neglect prognosis.

What we can state here with certainty is that the different ways to measure spatial neglect and its chronicity used in different studies contributes to the heterogeneity of the results reported in previous prognostic studies. Another factor contributing to this heterogeneity between prognostic studies is the fact that control patients are sometimes included and sometimes not. Inconsistent statements exist in the literature about whether control patients should be included. One should have in mind that predictions of chronic behavior of control patients do very likely boost the performance due to the absence of the target deficit from stroke onset. In general, consideration of control patients when calculating model accuracy should depend on the underlying research question. The algorithm of the present study was designed to minimize the prediction error for chronic and recovered neglect patients only, as we asked which features can distinguish chronic versus recovered patients.

### Limitations

In the present study, we used PCA to generate useful spatial features, considering the predictive power, feature space, and computational resources. But at the same time, principal components are less clinically applicable because their prognostic information cannot be used directly, as it needs to be translated to an individual patient first. Nevertheless, predictive components can be used to identify relevant lesion locations that might aid the development of future lesion predictors. In addition, we utilized a repeated nested CV approach to overcome overfitting. The out-of-sample predictions still relied on the same patient cohort as the model building, with respect to the preprocessing pipeline of the imaging data, imaging parameters, and neuropsychological tests. Therefore, external validation is needed to underpin the here presented findings (see Hope et al., 2023).

Other variables have been examined in the literature to predict neglect recovery (for review, see Durfee & Hillis, 2023) that were not used in the current study. These include, for example, premorbid brain atrophy (Levine et al., 1986), burden of white matter hyperintensities (Kamakura et al., 2017), fractional anisotropy values (Lunven et al., 2015), anosognosia (Stone et al., 1992), acute visual field defects (Samuelsson et al., 1997), acute allocentric (but not egocentric) neglect severity (Moore et al., 2021), as well as activation patterns and functional connectivity (Cao et al., 2022; Umarova et al., 2016). Changes in structural connectivity might also play a role (see Talozzi et al., 2023), although, for post-stroke aphasia, this measure could not add predictive value to models using the lesion information itself (Halai et al., 2020; Zhao et al., 2023). Future studies should investigate whether (some of) these or other potentially prognostic factors can add predictive information to the best-performing model(s) of the current study.

## Conclusion

Among different measures of long-term neglect prognosis, the direct improvement between acute and chronic stages of stroke was best predicted by using acute neglect severity as well as location and size of individual lesion maps overlapping with a previously reported chronic neglect ROI (Karnath et al., 2011). This model has achieved a remarkably high level of prediction accuracy, with 66% of the behavioral variance explained. Demographic data were not informative for the algorithm, whereas the initial behavior and acute lesion location emerged as useful predictors for individual neglect prognoses. Therefore, it is worthwhile to determine topographical features of a brain lesion when predicting chronicity of neglect. All in all, a prediction accuracy of almost two-thirds explained variance has great potential to help guide individualized therapeutic approaches to treat spatial neglect in the future. Clinicians could use this information to begin rehabilitation of neglect earlier and provide more intensive or frequent therapy sessions to prevent persistent neglect in patients predicted to become chronic.

## Data Availability

Patient-specific data cannot be made publicly available due to data protection restrictions. Supporting results including the predictive lesion location maps are openly available at Mendeley Data (DOI: 10.17632/njxw52k5sm.1).

https://data.mendeley.com/datasets/njxw52k5sm/1

## Acknowledgments

This work was supported by the Deutsche Forschungsgemeinschaft (KA 1258/23-1). Daniel Wiesen was supported by the Luxembourg National Research Fund (FNR/11601161). We thank Hannah Rosenzopf for methodological discussion.

## Data availability statement

Patient-specific data cannot be made publicly available due to data protection restrictions. Supporting results including the predictive lesion location maps are openly available at Mendeley Data (DOI: 10.17632/njxw52k5sm.1; https://data.mendeley.com/datasets/njxw52k5sm/1).

## Abbreviations

*CoC*: center of cancellation
*CV*: cross validation
*FS*: feature selection
*IPL*: inferior parietal lobule
*MSE*: mean squared error
*MTG/STG*: middle/superior temporal gyrus
*PC(A)*: principal component (analysis)
*ROI*: region of interest
*R²*: coefficient of determination
*SLF*: superior longitudinal fasciculus
*SVR*: support vector regression
*WB*: whole-brain.

## References

Arlot, S., & Celisse, A. (2010). A survey of cross-validation procedures for model selection. Statistics Surveys, 4(none), 40–79. 10.1214/09-SS054

Becker, E., & Karnath, H.-O. (2007). Incidence of visual extinction after left versus right hemisphere stroke. Stroke, 38(12), 3172–3174. 10.1161/STROKEAHA.107.489096

Brereton, R. G., & Lloyd, G. R. (2010). Support vector machines for classification and regression. The Analyst, 135(2), 230–267. 10.1039/b918972f

Cao, L., Ye, L., Xie, H., Zhang, Y., & Song, W. (2022). Neural substrates in patients with visual-spatial neglect recovering from right-hemispheric stroke. Frontiers in Neuroscience, 16. https://www.frontiersin.org/articles/10.3389/fnins.2022.974653

Chang, C.-C., & Lin, C.-J. (2011). LIBSVM: A library for support vector machines. ACM Transactions on Intelligent Systems and Technology, 2(3), 27:1-27:27. 10.1145/1961189.1961199

Chechlacz, M., Rotshtein, P., Roberts, K. L., Bickerton, W.-L., Lau, J. K. L., & Humphreys, G. W. (2012). The Prognosis of Allocentric and Egocentric Neglect: Evidence from Clinical Scans. PLOS ONE, 7(11), e47821. 10.1371/journal.pone.0047821

Clas, P., Groeschel, S., & Wilke, M. (2012). A Semi-Automatic Algorithm for Determining the Demyelination Load in Metachromatic Leukodystrophy. Academic Radiology, 19(1), 26–34. 10.1016/j.acra.2011.09.008

de Haan, B., Clas, P., Juenger, H., Wilke, M., & Karnath, H.-O. (2015). Fast semi-automated lesion demarcation in stroke. NeuroImage. Clinical, 9, 69–74. 10.1016/j.nicl.2015.06.013

Di Monaco, M., Schintu, S., Dotta, M., Barba, S., Tappero, R., & Gindri, P. (2011). Severity of Unilateral Spatial Neglect Is an Independent Predictor of Functional Outcome After Acute Inpatient Rehabilitation in Individuals With Right Hemispheric Stroke. Archives of Physical Medicine and Rehabilitation, 92(8), 1250–1256. 10.1016/j.apmr.2011.03.018

Durfee, A. Z., & Hillis, A. E. (2023). Unilateral Spatial Neglect Recovery Poststroke. Stroke, 54(1), 10–19. 10.1161/STROKEAHA.122.041710

Fan, L., Li, H., Zhuo, J., Zhang, Y., Wang, J., Chen, L., Yang, Z., Chu, C., Xie, S., Laird, A. R., Fox, P. T., Eickhoff, S. B., Yu, C., & Jiang, T. (2016). The Human Brainnetome Atlas: A New Brain Atlas Based on Connectional Architecture. Cerebral Cortex, 26(8), 3508–3526. 10.1093/cercor/bhw157

Farnè, A., Buxbaum, L. J., Ferraro, M., Frassinetti, F., Whyte, J., Veramonti, T., Angeli, V., Coslett, H. B., & Làdavas, E. (2004). Patterns of spontaneous recovery of neglect and associated disorders in acute right brain-damaged patients. Journal of Neurology, Neurosurgery, and Psychiatry, 75(10), 1401–1410. 10.1136/jnnp.2002.003095

Gauthier, L., Dehaut, F., & Joanette, Y. (1989). The Bells Test: A quantitative and qualitative test for visual neglect. International Journal of Clinical Neuropsychology, 11, 49–54.

Grasso, M. G., Troisi, E., Rizzi, F., Morelli, D., & Paolucci, S. (2005). Prognostic factors in multidisciplinary rehabilitation treatment in multiple sclerosis: An outcome study. Multiple Sclerosis Journal, 11(6), 719–724. 10.1191/1352458505ms1226oa

Halai, A. D., Woollams, A. M., & Lambon Ralph, M. A. (2020). Investigating the effect of changing parameters when building prediction models for post-stroke aphasia. Nature Human Behaviour, 4(7), Article 7. 10.1038/s41562-020-0854-5

Hier, D. B., Mondlock, J., & Caplan, L. R. (1983). Recovery of behavioral abnormalities after right hemisphere stroke. Neurology, 33(3), 345–345. 10.1212/WNL.33.3.345

Hillis, A. E., Beh, Y. Y., Sebastian, R., Breining, B., Tippett, D. C., Wright, A., Saxena, S., Rorden, C., Bonilha, L., Basilakos, A., Yourganov, G., & Fridriksson, J. (2018). Predicting recovery in acute poststroke aphasia. Annals of Neurology, 83(3), 612–622. 10.1002/ana.25184

Hope, T. M. H., Leff, A. P., & Price, C. J. (2018). Predicting language outcomes after stroke: Is structural disconnection a useful predictor? NeuroImage. Clinical, 19, 22–29. 10.1016/j.nicl.2018.03.037

Hope, T. M. H., Neville, D., Talozzi, L., Foulon, C., Forkel, S. J., Thiebaut de Schotten, M., & Price, C. J. (2023). Testing the Disconnectome Symptom Discoverer model on out-of-sample post-stroke language outcomes. *Brain*, awad352. 10.1093/brain/awad352

Hope, T. M. H., Seghier, M. L., Leff, A. P., & Price, C. J. (2013). Predicting outcome and recovery after stroke with lesions extracted from MRI images. NeuroImage: Clinical, 2, 424–433. 10.1016/j.nicl.2013.03.005

Imura, T., Mitsutake, T., Hori, T., & Tanaka, R. (2022). Predicting the prognosis of unilateral spatial neglect using magnetic resonance imaging in patients with stroke: A systematic review. Brain Research, 1789, 147954. 10.1016/j.brainres.2022.147954

Iosa, M., Morone, G., Antonucci, G., & Paolucci, S. (2021). Prognostic Factors in Neurorehabilitation of Stroke: A Comparison among Regression, Neural Network, and Cluster Analyses. Brain Sciences, 11(9), Article 9. 10.3390/brainsci11091147

Jehkonen, M., Ahonen, J.-P., Dastidar, P., Koivisto, A.-M., Laippala, P., Vilkki, J., & Molnár, G. (2000). Visual neglect as a predictor of functional outcome one year after stroke. Acta Neurologica Scandinavica, 101(3), 195–201. 10.1034/j.1600-0404.2000.101003195.x

Jehkonen, M., Laihosalo, M., Koivisto, A.-M., Dastidar, P., & Ahonen, J.-P. (2007). Fluctuation in Spontaneous Recovery of Left Visual Neglect: A 1-Year Follow-Up. European Neurology, 58(4), 210–214. 10.1159/000107941

Johannsen, L., & Karnath, H.-O. (2004). How Efficient is a Simple Copying Task to Diagnose Spatial Neglect in its Chronic Phase? Journal of Clinical and Experimental Neuropsychology, 26(2), 251–256. 10.1076/jcen.26.2.251.28085

Kalra, L., Perez, I., Gupta, S., & Wittink, M. (1997). The Influence of Visual Neglect on Stroke Rehabilitation. Stroke, 28(7), 1386–1391. 10.1161/01.STR.28.7.1386

Kamakura, C. K., Ueno, Y., Sakai, Y., Yoshida, H., Aiba, S., Hayashi, A., Shimura, H., Takeda, K., Kamakura, K., Hattori, N., & Urabe, T. (2017). White matter lesions and cognitive impairment may be related to recovery from unilateral spatial neglect after stroke. Journal of the Neurological Sciences, 379, 241–246. 10.1016/j.jns.2017.06.021

Karnath, H., Himmelbach, M., & Rorden, C. (2002). The subcortical anatomy of human spatial neglect: Putamen, caudate nucleus and pulvinar. Brain, 125(2), 350–360. 10.1093/brain/awf032

Karnath, H.-O., Rennig, J., Johannsen, L., & Rorden, C. (2011). The anatomy underlying acute versus chronic spatial neglect: A longitudinal study. Brain: A Journal of Neurology, 134(Pt 3), 903–912. 10.1093/brain/awq355

Karnath, H.-O., Rorden, C., & Ticini, L. F. (2009). Damage to White Matter Fiber Tracts in Acute Spatial Neglect. Cerebral Cortex, 19(10), 2331–2337. 10.1093/cercor/bhn250

Kasties, V., Karnath, H.-O., & Sperber, C. (2021). Strategies for feature extraction from structural brain imaging in lesion-deficit modelling. Human Brain Mapping, 42(16), 5409–5422. 10.1002/hbm.25629

Krstajic, D., Buturovic, L. J., Leahy, D. E., & Thomas, S. (2014). Cross-validation pitfalls when selecting and assessing regression and classification models. Journal of Cheminformatics, 6(1), 10. 10.1186/1758-2946-6-10

Levine, D. N., Warach, J. D., Benowitz, L., & Calvanio, R. (1986). Left Spatial neglect: Effects of lesion size and premorbid brain atrophy on severity and recovery following right cerebral infarction. Neurology, 36(3), 362–362. 10.1212/WNL.36.3.362

Lunven, M., Thiebaut De Schotten, M., Bourlon, C., Duret, C., Migliaccio, R., Rode, G., & Bartolomeo, P. (2015). White matter lesional predictors of chronic visual neglect: A longitudinal study. Brain: A Journal of Neurology, 138(Pt 3), 746–760. 10.1093/brain/awu389

Moore, M. J., Vancleef, K., Riddoch, M. J., Gillebert, C. R., & Demeyere, N. (2021). Recovery of Visuospatial Neglect Subtypes and Relationship to Functional Outcome Six Months After Stroke. Neurorehabilitation and Neural Repair, 35(9), 823–835. 10.1177/15459683211032977

Mori, S., Oishi, K., Jiang, H., Jiang, L., Li, X., Akhter, K., Hua, K., Faria, A. V., Mahmood, A., Woods, R., Toga, A. W., Pike, G. B., Neto, P. R., Evans, A., Zhang, J., Huang, H., Miller, M. I., van Zijl, P., & Mazziotta, J. (2008). Stereotaxic white matter atlas based on diffusion tensor imaging in an ICBM template. NeuroImage, 40(2), 570–582. 10.1016/j.neuroimage.2007.12.035

Rengachary, J., He, B., Shulman, G., & Corbetta, M. (2011). A Behavioral Analysis of Spatial Neglect and its Recovery After Stroke. Frontiers in Human Neuroscience, 5. https://www.frontiersin.org/articles/10.3389/fnhum.2011.00029

Röhrig, L., Sperber, C., Bonilha, L., Rorden, C., & Karnath, H.-O. (2022). Right hemispheric white matter hyperintensities improve the prediction of spatial neglect severity in acute stroke. NeuroImage: Clinical, 36, 103265. 10.1016/j.nicl.2022.103265

Rorden, C., Bonilha, L., Fridriksson, J., Bender, B., & Karnath, H.-O. (2012). Age-specific CT and MRI templates for spatial normalization. NeuroImage, 61(4), 957–965. 10.1016/j.neuroimage.2012.03.020

Rorden, C., & Karnath, H.-O. (2004). Using human brain lesions to infer function: A relic from a past era in the fMRI age? Nature Reviews. Neuroscience, 5(10), 813–819. 10.1038/nrn1521

Rorden, C., & Karnath, H.-O. (2010). A simple measure of neglect severity. Neuropsychologia, 48(9), 2758–2763. 10.1016/j.neuropsychologia.2010.04.018

Rorden, C., Karnath, H.-O., & Bonilha, L. (2007). Improving Lesion-Symptom Mapping. Journal of Cognitive Neuroscience, 19(7), 1081–1088. 10.1162/jocn.2007.19.7.1081

Saj, A., Verdon, V., Vocat, R., & Vuilleumier, P. (2012). ‘The anatomy underlying acute versus chronic spatial neglect’ also depends on clinical tests. Brain, 135(2), e207. 10.1093/brain/awr227

Samuelsson, H., Jensen, C., Ekholm, S., Naver, H., & Blomstrand, C. (1997). Anatomical and Neurological Correlates of Acute and Chronic Visuospatial Neglect Following Right Hemisphere Stroke* *Some of the results of this study were presented at: The 5th Nordic Meeting in Neuropsychology, Uppsala, Sweden, August 17–19, 1995. *Cortex*, *33*(2), 271–285. 10.1016/S0010-9452(08)70004-2

Shah, S., Vanclay, F., & Cooper, B. (1990). Efficiency, effectiveness, and duration of stroke rehabilitation. Stroke, 21(2), 241–246. 10.1161/01.STR.21.2.241

Smola, A. J., & Schölkopf, B. (2004). A tutorial on support vector regression. Statistics and Computing, 14(3), 199–222. 10.1023/B:STCO.0000035301.49549.88

Sperber, C., Gallucci, L., Mirman, D., Arnold, M., & Umarova, R. M. (2023). Stroke lesion size – Still a useful biomarker for stroke severity and outcome in times of high-dimensional models. NeuroImage: Clinical, 40, 103511. 10.1016/j.nicl.2023.103511

Stone, S. P., Patel, P., Greenwood, R. J., & Halligan, P. W. (1992). Measuring visual neglect in acute stroke and predicting its recovery: The visual neglect recovery index. Journal of Neurology, Neurosurgery, and Psychiatry, 55(6), 431–436.

Talozzi, L., Forkel, S. J., Pacella, V., Nozais, V., Allart, E., Piscicelli, C., Pérennou, D., Tranel, D., Boes, A., Corbetta, M., Nachev, P., & Thiebaut de Schotten, M. (2023). Latent disconnectome prediction of long-term cognitive-behavioural symptoms in stroke. Brain, 146(5), 1963–1978. 10.1093/brain/awad013

Thiebaut de Schotten, M., Tomaiuolo, F., Aiello, M., Merola, S., Silvetti, M., Lecce, F., Bartolomeo, P., & Doricchi, F. (2014). Damage to White Matter Pathways in Subacute and Chronic Spatial Neglect: A Group Study and 2 Single-Case Studies with Complete Virtual “In Vivo” Tractography Dissection. Cerebral Cortex, 24(3), 691–706. 10.1093/cercor/bhs351

Umarova, R. M., Nitschke, K., Kaller, C. P., Klöppel, S., Beume, L., Mader, I., Martin, M., Hennig, J., & Weiller, C. (2016). Predictors and signatures of recovery from neglect in acute stroke. Annals of Neurology, 79(4), 673–686. 10.1002/ana.24614

Varoquaux, G., Raamana, P. R., Engemann, D. A., Hoyos-Idrobo, A., Schwartz, Y., & Thirion, B. (2017). Assessing and tuning brain decoders: Cross-validation, caveats, and guidelines. NeuroImage, 145, 166–179. 10.1016/j.neuroimage.2016.10.038

Weintraub, S., & Mesulam, M. M. (1985). Mental state assessment of the young and elderly adults in behavioral neurology. Principles of Behavioral Neurology, 71–123.

Zhang, Y., Kimberg, D. Y., Coslett, H. B., Schwartz, M. F., & Wang, Z. (2014). Multivariate lesion-symptom mapping using support vector regression. Human Brain Mapping, 35(12), 5861– 5876. 10.1002/hbm.22590

Zhao, Y., Cox, C. R., Lambon Ralph, M. A., & Halai, A. D. (2023). Using in vivo functional and structural connectivity to predict chronic stroke aphasia deficits. Brain, 146(5), 1950–1962. 10.1093/brain/awac388

