## Supplement for "Predicting individual long-term prognosis of spatial neglect based on acute stroke patient data"

### Supplementary Methods

*Target variables*

Calculation of recovery:

We calculated the effectiveness of recovery based on a previously reported formula (adapted from Grasso et al., 2005; Shah et al., 1990), in which we used the value zero for the “maximum score” since a CoC or copying error of zero represents no deficit. Note that a recovery of greater than 100% was set to 100% before averaging. The formula was as follows:

$$recovery = \left(\frac{chronic\ score - acute\ score}{0 - acute\ score}\right) * 100\%$$

*Predictor variables*

Selection of principal components:

For each target variable and map variant separately, we tried different thresholds of cumulative explained variance and used the winning threshold (i.e., that produced the most accurate predictions) for further analyses (*chronic z-score*: WB lesion map 98% [N = 50], ROI overlap 90% [N = 19]; *difference*: WB lesion map 98% [N = 50], ROI overlap 100% [N = 71]; *recovery*: WB lesion map 100% [N = 71], ROI overlap 100% [N = 71]).

Again, for each target variable and map variant separately, we tried three different feature selection filter methods (minimum redundancy maximum relevance algorithm [MRMR], univariate feature ranking for regression using F-tests [F-test], neighborhood component analysis [NCA]). We then selected the five most important components that were most strongly associated with the target variable, identified by the winning filter method (*chronic z-score*: WB lesion map – F-test [PCs 13, 5, 36, 29, 55], ROI overlap – NCA [PCs 14, 13, 1, 2, 38]; *difference*: WB lesion map – NCA [PCs 3, 7, 26, 10, 11], ROI overlap – F-test [PCs 14, 58, 48, 2, 13]; *recovery*: WB lesion map – NCA [PCs 3, 1, 2, 6, 55], ROI overlap – F-test [PCs 50, 62, 25, 18, 4]).

### Supplementary Figures

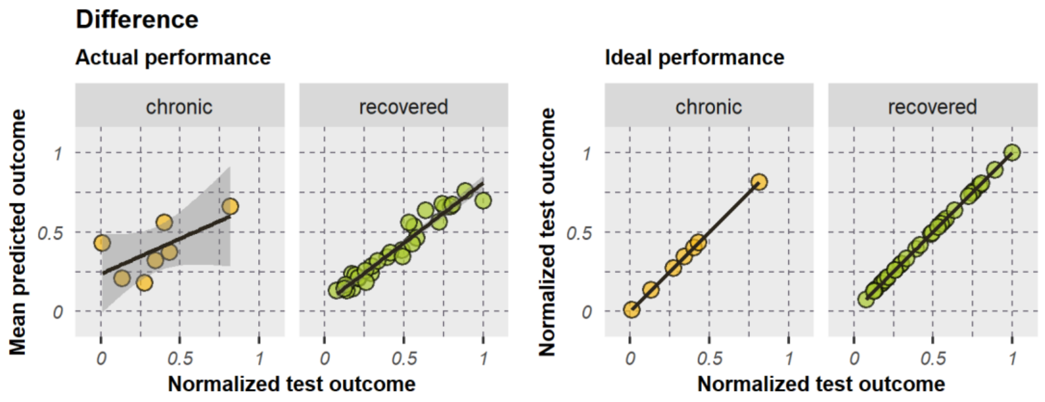


**Figure S1. Model fit of the best predictive model.** Model performance is illustrated for chronic (*N* = 7) and recovered patients (*N* = 30). Chronic neglect patients (*N* = 5), who were also included in the creation of the chronic neglect ROI (Karnath et al., 2011), were excluded (compare with Fig. 5C in the main manuscript).

### Supplementary Tables

**Table S1. Model performances for predicting the chronic z-score.**

| Model | Predictor(s) | MSE | $R^2$ | r | *p*(r) |
|---|---|---|---|---|---|
| Single/FS-1 | Acute behavior | 0.0543 | -0.101 | -0.139 | n.s. |
| | **Age** | **0.0490** | **0.007** | **0.115** | **n.s.** |
| | Sex | 0.0501 | -0.015 | -0.539 | *** |
| | WB lesion size | 0.0505 | -0.025 | 0.036 | n.s. |
| | **ROI lesion size** | **0.0372** | **0.246** | **0.525** | ***** |
| | WB-%-PCs | 0.0517 | -0.048 | -0.542 | *** |
| | **WB-FS-PCs** | **0.0223** | **0.548** | **0.771** | ***** |
| | ROI-%-PCs | 0.0549 | -0.114 | -0.021 | n.s. |
| | **ROI-FS-PCs** | **0.0460** | **0.068** | **0.271** | **n.s.** |
| FS-2 | Acute behavior | 0.0418 | 0.153 | 0.410 | ** |
| | Age | 0.0305 | 0.381 | 0.642 | *** |
| | Sex | 0.0400 | 0.189 | 0.473 | ** |
| | WB lesion size | 0.0470 | 0.047 | 0.274 | n.s. |
| | ROI lesion size | 0.0360 | 0.271 | 0.563 | *** |
| Full model | WB-%-PCs | 0.0517 | -0.048 | -0.153 | n.s. |
| | **WB-FS-PCs** | **0.0465** | **0.058** | **0.263** | **n.s.** |
| | ROI-%-PCs | 0.0514 | -0.041 | -0.063 | n.s. |
| | ROI-FS-PCs | 0.0554 | -0.124 | -0.069 | n.s. |

*Note*. Prediction accuracies are presented for models predicting the chronic z-score (chronic neglect severity). Performance was measured using the mean squared error (MSE), coefficient of determination ($R^2$), and Pearson correlation coefficient (r, with its corresponding significance value *p*; n.s. – not significant, * – $p < 0.05$, ** – $p < 0.01$, *** – $p < 0.001$). Models used either single predictors (“single”), combinations of thereof by applying forward sequential feature selection (“FS-1” represents the first iteration, “FS-2” represents the second iteration testing the winning predictor of the first iteration and a second variable), and full models using all variables but only one lesion location variable. Results of predictive variables (that explain some variance or improve the model) are in bold type, whereas the row of the selected predictor (i.e., winning model) is highlighted in grey. Note that an ideal model would achieve values of $MSE = 0$, $R^2 = 1$, $r = 1$ (a negative $R^2$ and a negative r represents a model with a very poor model fit).
Abbreviations: WB – whole-brain, ROI – (chronic neglect) region of interest, % – PCs selected according to a certain proportion of cumulatively explained variance, FS – PCs identified by a filter feature selection approach (five most important PCs selected), PCs – principal components.

**Table S2. Model performances for predicting the z-score difference.**

| Model | Predictor(s) | MSE | $R^2$ | r | *p*(r) |
|---|---|---|---|---|---|
| Single/FS-1 | **Acute behavior** | **0.0263** | **0.588** | **0.768** | ******* |
| | Age | 0.1170 | -0.830 | -0.085 | n.s. |
| | Sex | 0.1303 | -1.037 | 0.007 | n.s. |
| | WB lesion size | 0.0853 | -0.335 | 0.231 | n.s. |
| | ROI lesion size | 0.1522 | -1.380 | -0.015 | n.s. |
| | WB-%-PCs | 0.0693 | -0.084 | 0.310 | * |
| | WB-FS-PCs | 0.0797 | -0.247 | 0.294 | n.s. |
| | ROI-%-PCs | 0.0772 | -0.207 | 0.161 | n.s. |
| | ROI-FS-PCs | 0.0960 | -0.501 | 0.230 | n.s. |
| FS-2 | **Age** | **0.0251** | **0.608** | **0.780** | ******* |
| | Sex | 0.0283 | 0.557 | 0.747 | *** |
| | WB lesion size | 0.0267 | 0.582 | 0.763 | *** |
| | **ROI lesion size** | **0.0239** | **0.626** | **0.792** | ******* |
| | WB-%-PCs | 0.0278 | 0.566 | 0.753 | *** |
| | **WB-FS-PCs** | **0.0262** | **0.590** | **0.769** | ******* |
| | **ROI-%-PCs** | **0.0251** | **0.607** | **0.784** | ******* |
| | **ROI-FS-PCs** | **0.0223** | **0.651** | **0.807** | ******* |
| FS-3 | Age | 0.0257 | 0.598 | 0.776 | *** |
| | Sex | 0.0254 | 0.602 | 0.777 | *** |
| | **WB lesion size** | **0.0221** | **0.655** | **0.811** | ******* |
| | **ROI lesion size** | **0.0216** | **0.662** | **0.814** | ******* |
| FS-4 | Age | 0.0235 | 0.632 | 0.797 | *** |
| | Sex | 0.0250 | 0.609 | 0.781 | *** |
| | WB lesion size | 0.0254 | 0.603 | 0.777 | *** |
| Full model | **WB-%-PCs** | **0.0301** | **0.530** | **0.728** | ******* |
| | **WB-FS-PCs** | **0.0299** | **0.533** | **0.731** | ******* |
| | **ROI-%-PCs** | **0.0289** | **0.548** | **0.741** | ******* |
| | **ROI-FS-PCs** | **0.0280** | **0.563** | **0.752** | ******* |

*Note*. Prediction accuracies are presented for models predicting the chronic z-score (chronic neglect severity). Performance was measured using the mean squared error (MSE), coefficient of determination ($R^2$), and Pearson correlation coefficient (r, with its corresponding significance value *p*; n.s. – not significant, * – $p < 0.05$, ** – $p < 0.01$, *** – $p < 0.001$). Models used either single predictors (“single”), combinations of thereof by applying forward sequential feature selection (“FS-1” represents the first iteration, “FS-2” represents the second iteration testing the winning predictor of FS-1 and a second variable, “FS-3” represents the third iteration testing the winning combination of FS-2 and a third variable, and so on), and full models using all variables but only one lesion location variable. Results of predictive variables (that explain some variance or improve the model) are in bold type, whereas rows of the selected predictors (i.e., winning models) are highlighted in grey. Note that an ideal model would achieve values of $MSE = 0$, $R^2 = 1$, $r = 1$ (a negative $R^2$ and a negative r represents a model with a very poor model fit).
*Abbreviations*: WB – whole-brain, ROI – (chronic neglect) region of interest, % – PCs selected according to a certain proportion of cumulatively explained variance, FS – PCs identified by a filter feature selection approach (five most important PCs selected), PCs – principal components.

**Table S3. Model performances for predicting the effectiveness of recovery.**

| Model | Predictor(s) | MSE | $R^2$ | r | *p*(r) |
|---|---|---|---|---|---|
| Single/FS-1 | Acute behavior | 0.0698 | -0.120 | -0.354 | * |
| | Age | 0.0676 | -0.084 | 0.223 | n.s. |
| | Sex | 0.0680 | -0.090 | -0.046 | n.s. |
| | WB lesion size | 0.0662 | -0.062 | 0.096 | n.s. |
| | ROI lesion size | 0.0857 | -0.374 | 0.265 | n.s. |
| | WB-%-PCs | 0.0673 | -0.079 | -0.393 | * |
| | WB-FS-PCs | 0.0685 | -0.098 | -0.144 | n.s. |
| | ROI-%-PCs | 0.0687 | -0.102 | -0.169 | n.s. |
| | ROI-FS-PCs | 0.0665 | -0.067 | 0.124 | n.s. |
| FS-2 | Acute behavior | 0.0701 | -0.123 | -0.310 | * |
| | Age | 0.0687 | -0.101 | -0.095 | n.s. |
| | Sex | 0.0699 | -0.121 | -0.136 | n.s. |
| | ROI lesion size | 0.0704 | -0.129 | 0.002 | n.s. |
| | WB-%-PCs | 0.0685 | -0.098 | -0.395 | ** |
| | WB-FS-PCs | 0.0695 | -0.115 | -0.182 | n.s. |
| | ROI-%-PCs | 0.0699 | -0.120 | -0.274 | n.s. |
| | ROI-FS-PCs | 0.0688 | -0.104 | 0.020 | n.s. |
| Full model | WB-%-PCs | 0.0710 | -0.139 | -0.431 | ** |
| | WB-FS-PCs | 0.0732 | -0.174 | -0.452 | ** |
| | ROI-%-PCs | 0.0684 | -0.096 | -0.311 | * |
| | ROI-FS-PCs | 0.0706 | -0.132 | -0.202 | n.s. |

*Note*. Prediction accuracies are presented for models predicting the chronic z-score (chronic neglect severity). Performance was measured using the mean squared error (MSE), coefficient of determination ($R^2$), and Pearson correlation coefficient (r, with its corresponding significance value *p*; n.s. – not significant, * – $p < 0.05$, ** – $p < 0.01$, *** – $p < 0.001$). Models used either single predictors (“single”), combinations of thereof by applying forward sequential feature selection (“FS-1” represents the first iteration, “FS-2” represents the second iteration testing the winning predictor of the first iteration and a second variable), and full models using all variables but only one lesion location variable. Note that none of the models could explain some variance. The row of the selected predictor (i.e., winning model) is highlighted in grey. Note that an ideal model would achieve values of $MSE = 0$, $R^2 = 1$, $r = 1$ (a negative $R^2$ and a negative r represents a model with a very poor model fit).
*Abbreviations*: WB – whole-brain, ROI – (chronic neglect) region of interest, % – PCs selected according to a certain proportion of cumulatively explained variance, FS – PCs identified by a filter feature selection approach (five most important PCs selected), PCs – principal components.

**Table S4. Brain regions and tracts included in principal components predictive for neglect prognosis.**

| Atlas | Components of whole-brain lesion maps | | | Components of ROI-based overlaps | | |
|---|---|---|---|---|---|---|
| | Gyrus/Tract | Label | *N* | Gyrus/Tract | Label | *N* |
| Gray matter (BNA) | MTG | 88 | 4248 | MTG | 88 | 2524 |
| | Putamen | 230 | 2061 | Putamen | 230 | 2106 |
| | Insular Gyrus | 164 | 1564 | STG | 80 | 1023 |
| | Insular Gyrus | 172 | 1488 | Globus Pallidus | 222 | 1013 |
| | Insular Gyrus | 174 | 1360 | Dorsal caudate | 228 | 627 |
| | Globus Pallidus | 222 | 1205 | IPL | 146 | 530 |
| | IPL | 146 | 1105 | STG | 76 | 517 |
| | STG | 80 | 1084 | Insular Gyrus | 170 | 322 |
| | Insular Gyrus | 170 | 820 | Putamen | 226 | 307 |
| | Dorsal caudate | 228 | 735 | STG | 78 | 81 |
| | Postcentral Gyrus | 158 | 716 | STG | 72 | 59 |
| | MTG | 82 | 696 | pSTS | 122 | 46 |
| | STG | 74 | 645 | MTG | 82 | 26 |
| | STG | 72 | 496 | Postcentral Gyrus | 158 | 12 |
| | pSTS | 122 | 443 | pSTS | 124 | 11 |
| | IPL | 136 | 292 | Nucleus accumbens | 224 | 11 |
| | Putamen | 226 | 289 | STG | 74 | 8 |
| | IPL | 138 | 261 | Insular Gyrus | 166 | 8 |
| | MTG | 86 | 229 | Insular Gyrus | 174 | 2 |
| | Insular Gyrus | 168 | 226 | Ventral caudate | 220 | 2 |
| | ITG | 100 | 211 | MTG | 84 | 1 |
| | LOcC | 210 | 133 | MTG | 86 | 1 |
| | LOcC | 202 | 117 | IPL | 142 | 1 |
| | STG | 76 | 100 | | | |
| | IFG | 38 | 74 | | | |
| | Precentral Gyrus | 62 | 70 | | | |
| | IPL | 140 | 63 | | | |
| | IPL | 144 | 60 | | | |
| | Precentral Gyrus | 54 | 47 | | | |
| | STG | 78 | 31 | | | |
| | Postcentral Gyrus | 160 | 25 | | | |
| | Postcentral Gyrus | 156 | 24 | | | |
| | Nucleus accumbens | 224 | 20 | | | |
| | IPL | 142 | 18 | | | |
| | Insular Gyrus | 166 | 16 | | | |
| | pSTS | 124 | 9 | | | |
| | ITG | 92 | 8 | | | |
| | MVOcC | 198 | 7 | | | |
| | LOcC | 200 | 6 | | | |
| | Amygdala | 214 | 5 | | | |
| | Precuneus | 152 | 3 | | | |
| | SPL | 130 | 2 | | | |
| | Ventral caudate | 220 | 1 | | | |
| White matter (JHU) | External capsule | 33 | 3066 | External capsule | 33 | 1245 |
| | Superior corona radiata | 25 | 1764 | Internal capsule | 17 | 1156 |
| | SLF | 41 | 1479 | Posterior thalamic radiation | 29 | 332 |
| | Internal capsule | 17 | 1110 | SLF | 41 | 325 |
| | Posterior corona radiata | 27 | 612 | Internal capsule | 21 | 285 |
| | Internal capsule | 21 | 501 | Internal capsule | 19 | 164 |
| | Internal capsule | 19 | 461 | Superior corona radiata | 25 | 161 |
| | Anterior corona radiata | 23 | 457 | SFOF | 43 | 147 |

| | | | | | |
|---|---|---|---|---|---|
| SFOF | 43 | 306 | Posterior corona radiata | 27 | 49 |
| Posterior thalamic radiation | 29 | 185 | Uncinate fasciculus | 45 | 25 |
| Sagittal stratum | 31 | 25 | Sagittal stratum | 31 | 4 |
| Uncinate fasciculus | 45 | 17 | | | |
| Tapetum | 47 | 10 | | | |

*Note*. Table lists cortical brain regions and white matter tracts that were covered by the five most important principal components (preselected via feature selection filter) predictive for chronic neglect severity (left column, components derived from whole-brain lesion maps) or direct improvement from acute to chronic stage of stroke (right column, components derived from lesion maps overlapping with the chronic neglect ROI). Gray matter regions and their labels are based on the Brainnetome Atlas (BNA; Fan et al., 2016) with 246 parcels; white matter tracts and their labels are based on the JHU ICBM white matter atlas (Mori et al., 2008) with 48 tracts. Regions are sorted by decreasing number of overlapping 1mm isotropic voxels (*N*).

### References

Grasso, M. G., Troisi, E., Rizzi, F., Morelli, D., & Paolucci, S. (2005). Prognostic factors in multidisciplinary rehabilitation treatment in multiple sclerosis: An outcome study. Multiple Sclerosis Journal, 11(6), 719–724. https://doi.org/10.1191/1352458505ms1226oa

Fan, L., Li, H., Zhuo, J., Zhang, Y., Wang, J., Chen, L., Yang, Z., Chu, C., Xie, S., Laird, A. R., Fox, P. T., Eickhoff, S. B., Yu, C., & Jiang, T. (2016). The Human Brainnetome Atlas: A New Brain Atlas Based on Connectional Architecture. Cerebral Cortex, 26(8), 3508–3526. https://doi.org/10.1093/cercor/bhw157

Karnath, H.-O., Rennig, J., Johannsen, L., & Rorden, C. (2011). The anatomy underlying acute versus chronic spatial neglect: A longitudinal study. Brain: A Journal of Neurology, 134(Pt 3), 903–912. https://doi.org/10.1093/brain/awq355

Mori, S., Oishi, K., Jiang, H., Jiang, L., Li, X., Akhter, K., Hua, K., Faria, A. V., Mahmood, A., Woods, R., Toga, A. W., Pike, G. B., Neto, P. R., Evans, A., Zhang, J., Huang, H., Miller, M. I., van Zijl, P., & Mazziotta, J. (2008). Stereotaxic white matter atlas based on diffusion tensor imaging in an ICBM template. NeuroImage, 40(2), 570–582. https://doi.org/10.1016/j.neuroimage.2007.12.035

Shah, S., Vanclay, F., & Cooper, B. (1990). Efficiency, effectiveness, and duration of stroke rehabilitation. Stroke, 21(2), 241–246. https://doi.org/10.1161/01.STR.21.2.241
